# Impact of extended Elexacaftor/Tezacaftor/Ivacaftor therapy on the gut microbiome in cystic fibrosis

**DOI:** 10.1101/2024.02.01.24301982

**Authors:** Ryan Marsh, Claudio Dos Santos, Alexander Yule, Neele S Dellschaft, Caroline L Hoad, Christabella Ng, Giles Major, Alan R Smyth, Damian Rivett, Christopher van der Gast

## Abstract

**Background:** There is a paucity of knowledge on the longer-term effects of CF transmembrane conductance regulator (CFTR) modulator therapies upon the gut microbiome and associated outcomes. In a pilot study, we investigated longitudinal Elexacaftor/Tezacaftor/Ivacaftor (ETI) therapy on the gut microbiota, metabolomic functioning, and clinical outcomes in people with CF (pwCF).

**Study design:** Faecal samples from 20 pwCF were acquired before and then following 3, 6, and 17+ months of ETI therapy. Samples were subjected to microbiota sequencing and targeted metabolomics to profile and quantify short-chain fatty acid composition. Ten healthy matched controls were included for comparison. Clinical data, including markers of intestinal function were integrated to investigate relationships.

**Results:** Extended ETI therapy increased core microbiota diversity and composition, which translated to gradual shifts in whole microbiota composition towards that observed in healthy controls. Despite becoming more similar over time, CF microbiota and functional metabolite compositions remained significantly different to healthy controls. Antibiotic treatment for pulmonary infection significantly explained a relatively large degree of variation within the whole microbiota and rarer satellite taxa. Clinical outcomes were not significantly different following ETI.

**Conclusions:** A positive trajectory towards the microbiota observed in healthy controls was found. However, we posit that progression was predominately impeded by pulmonary antibiotics administration. We recommend future studies use integrated omics approaches within a combination of long-term longitudinal patient studies and model experimental systems. This will deepen our understanding of the impacts of CFTR modulator therapy and respiratory antibiotic interventions upon the gut microbiome and gastrointestinal pathophysiology in CF.

## 1. Introduction

Alongside the classical respiratory complications of cystic fibrosis (CF) are also the gastrointestinal abnormalities of disease. People with CF (pwCF) suffer from persistent intestinal symptoms and abnormalities that impact both morbidity and mortality [1]. This has continued despite the transition of treatment for CF entering the phase of highly effective CFTR modulator therapies, which correct the underlying defect of chloride and bicarbonate transport across epithelial surfaces of the body [2]. Although the majority of pwCF are now receiving CFTR modulator based therapy [3], the need to better understand the relationships between intestinal symptoms and abnormalities persists, as determined by pwCF, carers, and clinicians [4,5]. It is understood that intestinal dysbiosis, that is the disruption of the microbial communities inhabiting the intestinal tract, is frequent in CF and leads to compositions of bacteria distinctly different from healthy controls from birth and throughout adulthood [6–8]. Whilst this could be further compounded by the CF-associated lifestyle, such as dietary habits and frequent antibiotic usage [9,10], disruption of CFTR activity alone is seemingly sufficient to elicit structural changes to the microbiota [11]. Furthermore, the altered gut microbiota in pwCF has previously been associated with various manifestations of the GI tract [12–14]. Given this evidence, it is possible that CFTR modulators may alter the gut microbiota composition to resemble that of healthy individuals more closely, alongside improving other common intestinal abnormalities and symptoms in pwCF.

Previous studies investigating CFTR modulators and microbiota within the gastrointestinal tract have been mostly limited to Ivacaftor [15–18] and a couple of dual-modulator studies [17], including our previous work investigating the impact of Tezacaftor/Ivacaftor [19]. With respect to the microbiota structure, previous findings vary across studies incorporating different modulators, patient demographics, and treatment lengths [15–18]. Results for the frequently observed inflammation in the gut following modulator treatment are also contrasting. Some studies observed a reduction in faecal calprotectin following modulator usage [15,20,21], whilst others [16] and our own group [19], did not measure any notable reduction.

Following the approval of Elexacaftor/Tezacaftor/Ivacaftor (ETI) triple modulator therapy for pwCF from NHS England, over 60% of all pwCF in the United Kingdom are now registered users [3]. ETI therapy demonstrates increased clinical efficacy as compared to previous dual or mono-therapy approaches [22], yet little information is available on intestinal outcomes and the impact upon the gut microbiota. Given the scarcity of current evidence, the aim of this pilot study was to therefore assess the impact of ETI therapy in pwCF upon the intestinal microbiota. We characterised the respective gut microbiota of pancreatic insufficient pwCF ≥ 12 years, with at least one copy of the F508del mutation, before and during ETI therapy of up to 23 months duration. Furthermore, we combined microbiota data with targeted metabolomics of quantified faecal short-chain fatty acids (SCFAs), patient clinical data, and markers of intestinal function as measured by magnetic resonance imaging (MRI) [23]. Patient symptoms were also measured before and after initiating ETI therapy. Samples and clinical data from 10 matched healthy controls were available for analysis. We hypothesised that ETI therapy would increase gut microbiota diversity, reshape community structure, and potentially lead to functional changes to resemble outcomes measured in healthy controls.

## 2. Methods

### 2.1. Study participants and design

Twenty-four pwCF were initially recruited from Nottingham University Hospitals NHS Trust, with 20 participants ultimately providing at least one faecal sample following ETI therapy to form the cohort for our observational study. Participants attended clinic at baseline, 3, and 6 months following the initiation of ETI. A subset of pwCF (*n* = 7) also provided samples following extended ETI therapy of 19.8 ± 2.0 months (mean ± SD), with a minimum time of 17 months additional treatment. Additional faecal samples and metadata for 10 age and sex-matched healthy controls from our previous study were available for microbiota and metabolomic comparison also [24]. During study visits, participants provided faecal samples and completed magnetic resonance imaging (MRI) scans following the consumption of a standardised meal plan to detail their gut function as conducted previously [24], alongside specifying gut symptoms. This included the validated PAC-SYM questionnaire [25]. The full study design, including protocols, methods, and other results are described in the Supplementary Methods & Results. An overview of participant clinical characteristics is detailed in Table 1, with comprehensive participant information and MRI data detailed in Tables S1 and S2, respectively. Access to the MRI data (unpublished) was kindly granted by the University of Nottingham MRI research team at the Sir Peter Mansfield Imaging Centre. Written informed consent, or parental consent and assent for paediatric participants, was obtained from all participants. Study approval was obtained from the UK National Research Ethics Committee (20/PR/0508). All faecal samples obtained were immediately stored at - 80°C prior to processing for microbiota sequencing and metabolomics to reduce changes before downstream community analysis [26].

**Table 1.** Overall clinical characteristics of controls and pwCF at baseline.

| Characteristic | pwCF | Controls |
| --- | --- | --- |
| Baseline Age (Mean ± SD) | 21.0 ± 8.6 | 21.4 ± 7.4 |
| Male (%) | 15 (75) | 5 (50) |
| Baseline BMI (Mean ± SD) | 21.1 ± 3.4 | 22.9 ± 4.4 |
| F508del/F508del (%) | 13 (65) | 0 (0) |
| Baseline FEV1% (Mean ± SD) | 79.1 ± 20.5 | - |
| Pancreatic insufficient (%) | 20 (100) | 0 (0) |
| Cystic fibrosis-related diabetes (%) | 4 (20) | - |
| Antibiotics during study (%) | 14 (70) | 0 (0) |

### 2.2. Targeted amplicon sequencing

DNA from dead or damaged cells, as well as extracellular DNA was excluded from analysis via cross-linking with propidium monoazide (PMA) prior to DNA extraction, as previously described [27]. Cellular pellets resuspended in PBS were loaded into the ZYMO Quick-DNA Fecal/Soil Microbe Miniprep Kit (Cambridge Bioscience, Cambridge, UK) as per the manufacturer’s instructions. Dual mechanical-chemical sample disruption was performed using the FastPrep-24 5G instrument (MP Biomedicals, California, USA). Following DNA extraction, approximately 20 ng of template DNA was then amplified using Q5 high-fidelity DNA polymerase (New England Biolabs, Hitchin, UK) using a paired-end sequencing approach targeting the bacterial 16S rRNA gene region (V4-V5) as previously described [24]. Pooled barcoded amplicon libraries were sequenced on the Illumina MiSeq platform (V3 Chemistry). Extended methodology, primers and PCR conditions can be found in the Supplementary Methods & Results.

### 2.3. Sequence processing and analysis

Sequence processing and data analysis were then carried out in R (Version 4.2.1), utilising the packages DADA2 [28] and decontam [29]. The full protocol is detailed in the Supplementary Methods & Results. Raw sequence data reported in this study has been deposited in the European Nucleotide Archive under the study accession number PRJEB61286.

### 2.4 Gas-chromatography mass-spectrometry (GC-MS) of faecal samples to investigate SCFA levels

GC-MS analysis was carried out using an Agilent 7890B/5977 Single Quadrupole Mass Selective Detector (MSD) (Agilent Technologies) equipped with a non-polar HP-5ms Ultra Inert capillary column (30 m × 0.25 mm × 0.25 µm) (Agilent Technologies). Faecal sample processing to obtain, and then to derivatise SCFAs prior to GC-MS analysis, was carried out as previously described [19]. Following derivatisation, samples were loaded onto the GC-MS and injected using an Agilent 7693 Autosampler. MS grade water processed in parallel was used as a blank sample to correct the background. Selected ion monitoring (SIM) mode was used for subsequent analyses; all confirmation and target ions lists are summarised in Table S3. Agilent MassHunter workstation version B.07.00 programs were used to perform post-run analyses. A ^13^C-short chain fatty acids stool mixture (Merck Life Science, Poole, UK) was used as the internal standard to normalise all spectra obtained prior to analyses. A volatile fatty acid mixture (Merck Life Science, UK) was used to construct calibration curves for the quantification of target metabolites. Extended information surrounding sample processing, SCFA extraction, derivatisation, and GC-MS parameters can be found in the Supplementary Methods & Results.

### 2.5. Faecal Calprotectin measurement

Stool was extracted for downstream assays using the ScheBo® Master Quick-Prep (ScheBo Biotech, Giessen, Germany), according to the manufacturer instructions. Faecal calprotectin was analysed using the BÜHLMANN fCAL ELISA (Bühlmann Laboratories Aktiengesellschaft, Schonenbuch, Switzerland), according to the manufacturer’s protocol.

### 2.6. Statistical Analysis

Regression analysis, including calculated coefficients of determination (*r*^2^), degrees of freedom (df), *F*-statistic and significance values (*P*) were utilised for microbial partitioning into common core and rarer satellite groups, and were calculated using XLSTAT v2021.1.1 (Addinsoft, Paris, France). Fisher’s alpha index of diversity and the Bray-Curtis index of similarity were calculated using PAST v3.21 [30]. Tests for significant differences in microbiota diversity and SCFAs were performed using Kruskal-Wallis in XLSTAT. Student’s t-tests used to determine differences in metadata were also performed in XLSTAT. Analysis of similarities (ANOSIM) with Bonferroni correction was used to test for significance in microbiota and SCFA composition and was performed in PAST. Similarity of percentages (SIMPER) analysis, to determine which constituents drove compositional differences between groups, and was performed in PAST. Redundancy analysis (RDA), was performed in CANOCO v5 [31]. Statistical significance for all tests was deemed at the *P* ≤ 0.05 level.

## 3. Results

Following sequence processing and taxa assignment, the microbiota data across each pwCF treatment period from baseline, and the healthy control participants was partitioned for further sub-analysis. Microbiota data were partitioned into common abundant core taxa, and rarer less frequent satellite taxa following the establishment of significant distribution abundance relationships of the taxa across all treatment duration groups and the healthy control participants (Figure S1). The core taxa (Table S4) constituted 70.1 % of the total abundance for the healthy controls, whilst across pwCF they averaged 30.0% ± 6.1 (Mean ± SD).

Diversity of the whole microbiota, core taxa, and satellite taxa was plotted across the increasing ETI treatment periods in pwCF (Figure 1A). Overall, the diversity changes across all microbiota partitions included a gradual decline until 6 months of ETI treatment, highlighted by the significant reduction in diversity between baseline and 6 months for the whole microbiota (*P* = 0.001), core taxa (*P* = 0.003), and satellite taxa (*P* = 0.001) (Table S5). This was then succeeded by an increase in diversity following extended ETI treatment. The most pronounced change was the core taxa, for which diversity greatly increased following extended ETI treatment relative to other time points, reflected by significant increases in diversity compared to both baseline (P = 0.001) and the 6-month time period (P < 0.001) (Table S5). Despite the positive trajectory of diversity in pwCF following extended ETI therapy, there remained distinct differences when compared to healthy controls (Figure 1B, Table S6), whereby all partitions of the microbiota were significantly more diverse as compared to pwCF (*P* = 0.001).

**Figure 1.**
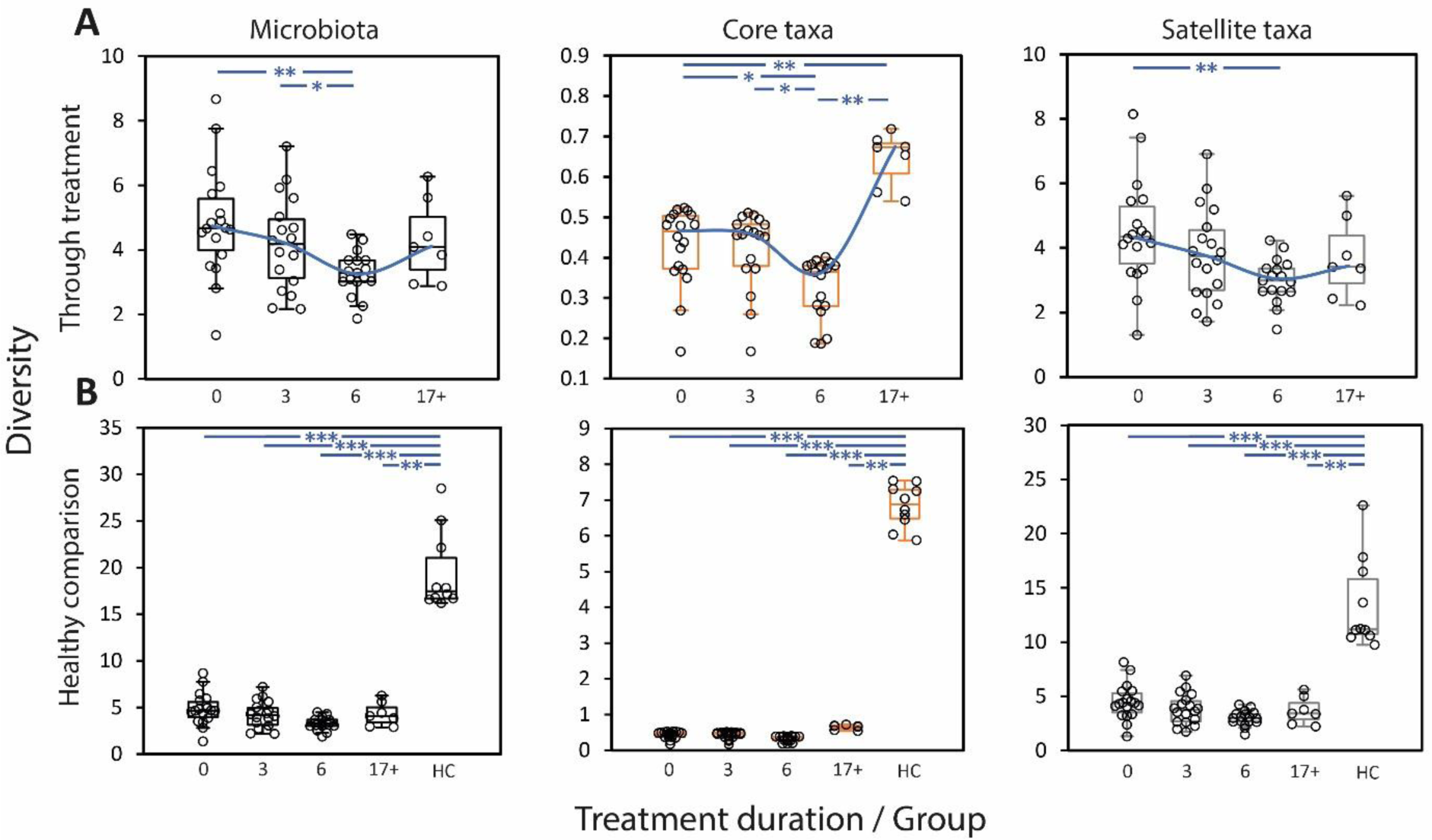
Microbiota diversity compared across groups utilising Fisher’s alpha index of diversity. Black plots indicate the whole microbiota, whilst orange and grey denote the core and satellite taxa respectively. (A) Diversity across pwCF at baseline or following ETI treatment. Black circles denote individual patient participant data. Error bars represent 1.5 times the inter-quartile range (IQR). Blue line indicates trend and direction of travel through the median of each treatment time point. Asterisks denote significant differences in diversity between treatment periods (sequential and to baseline) following Kruskal-Wallis testing (Table S5). (B) Diversity of pwCF and also healthy controls for comparison. Black circles denote individual patient participant data. Error bars represent 1.5 times the inter-quartile range (IQR). Asterisks denote significant differences in diversity between pwCF treatment periods and healthy controls following Kruskal-Wallis testing (Table S6). Abbreviations; 0,3 & 6 – Samping time point (months), 17+ – Extended (17+ months) sampling period, HC – Healthy control participants. *P* values; ***; P ≤ 0.0001, **; P ≤ 0.001, *; P ≤ 0.05.

When analysing microbiota composition throughout ETI treatment, similarity of the whole microbiota composition increased in pwCF as ETI treatment progressed, with a mean similarity (± SD) of 0.32 ± 0.09 following extended ETI (Figure 2A). Within-group similarity did however remain larger in the healthy controls, with a mean similarity of 0.44 ± 0.06. Within-group similarity for the satellite taxa initially increased but was relatively consistent throughout ETI treatment and was comparable to healthy controls following extended ETI (0.19 ± 0.04 and 0.19 ± 0.07, respectively). The core taxa similarity, however, increased in pwCF following extended ETI, surpassing that of healthy control participants (0.60 ± 0.12 and 0.57 ± 0.06, respectively). When comparing the composition of the microbiota between sequential ETI treatment periods (Figure 2B), the largest similarity across the whole microbiota and core taxa could be observed between 6 months and the extended ETI period, whilst the satellite taxa had no clear direction of travel. There were, however, significant differences between both the core (P < 0.001) and satellite (P = 0.035) taxa between months 3 and 6 of ETI treatment (Table S7). When comparisons were extended to include healthy controls (Figure 2C), all partitions of the microbiota in pwCF illustrated the highest resemblance to healthy controls following the extended ETI treatment period. Regardless of this increased similarity, microbiota composition of the healthy controls remained highly distinct and significantly different (*P* < 0.001) from pwCF throughout the study period (Figure 2C, Table S8).

**Figure 2.**
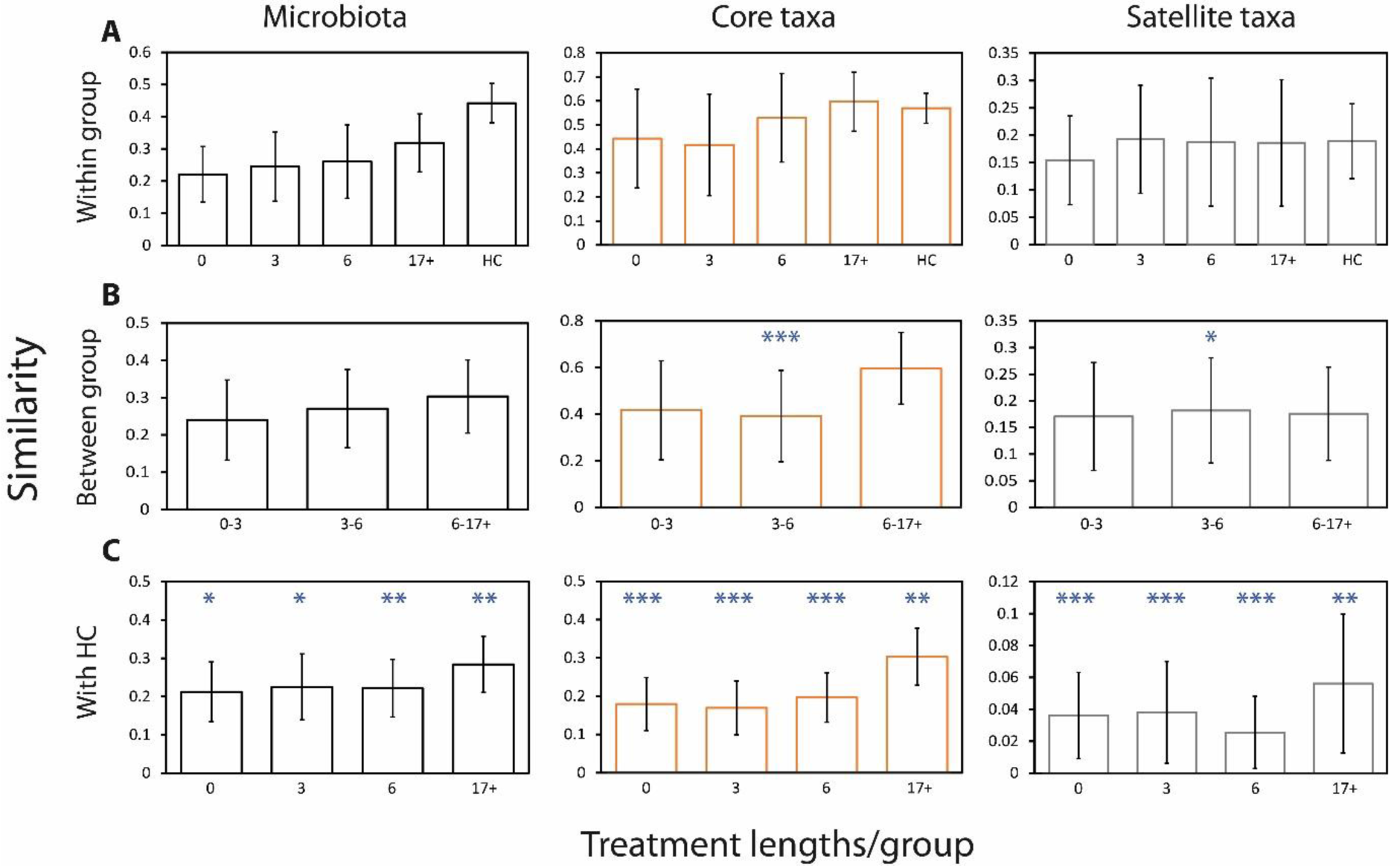
Microbiota similarity compared across groups utilising the Bray-Curtis index of similarity. (A) Within group similarity across pwCF at baseline or following ETI treatment, and healthy controls. (B) Between group similarity change over time in pwCF. (C) Similarity between pwCF on ETI at various time points and healthy control participants. Error bars represent standard deviation of the mean. Asterisks inicate significant differences in microbiota composition following the use of one-way ANOSIM testing. Summary statistics are presented in Tables S7-8. *P* values; ***; P ≤ 0.0001, **; P ≤ 0.001, *; P ≤ 0.05.

To understand which taxa were driving the dissimilarity maintained between healthy control participants and pwCF following extended ETI therapy, SIMPER analysis was conducted (Table 2). The top drivers of dissimilarity between the groups were *Blautia* sp. (OTU 1), *Bifidobacterium adolescentis*, *Bacteroides dorei*, *Eubacterium rectale*, and *Anaerostipes hadrus*. Overall, 24 taxa comprised over 50% of the dissimilarity between the groups and this consisted of many prominent SCFA producing bacteria from other genera, including *Faecalibacterium, Ruminococcus*, and *Collinsella,* Interestingly, some of these taxa were increased in the pwCF group compared to the healthy controls. Additional SIMPER analysis between baseline and following extended ETI therapy revealed that similar taxa were also drivers of the change seen across the ETI treatment period in pwCF (Table S9).

**Table 2.** Similarity of percentage (SIMPER) analysis of microbiota dissimilarity (Bray-Curtis) between healthy control and pwCF samples following 17+ months (extended) treatment with ETI.

| Taxa | % Relative abundance |  | Av. Dissimilarity | % Contribution | Cumulative (%) |
| --- | --- | --- | --- | --- | --- |
|  | 17+ Months | Healthy Controls |  |  |  |
| *Blautia 1* | 11.70 | 4.91 | 3.24 | 4.53 | 4.53 |
| *Bifidobacterium adolescentis* | 4.90 | 0.81 | 2.35 | 3.28 | 7.81 |
| *Bacteroides dorei* | 3.79 | 3.35 | 2.12 | 2.95 | 10.77 |
| *Eubacterium rectale* | 3.68 | 4.78 | 2.02 | 2.82 | 13.58 |
| *Anaerostipes hadrus* | 4.35 | 1.99 | 2.00 | 2.79 | 16.37 |
| *Fusicatenibacter saccharivorans* | 5.73 | 3.69 | 1.96 | 2.74 | 19.11 |
| *Ruminococcus bromii* | 3.34 | 2.93 | 1.93 | 2.70 | 21.81 |
| *Gemmiger formicilis* | 4.24 | 1.05 | 1.90 | 2.65 | 24.46 |
| *Blautia luti* | 5.55 | 3.01 | 1.63 | 2.27 | 26.73 |
| *Bacteroides vulgatus* | 1.66 | 3.32 | 1.62 | 2.26 | 28.99 |
| *Collinsella aerofaciens* | 3.33 | 0.19 | 1.59 | 2.23 | 31.21 |
| *Dorea longicatena* | 4.39 | 1.79 | 1.47 | 2.05 | 33.26 |
| *Eubacterium hallii* | 4.81 | 2.39 | 1.26 | 1.76 | 35.02 |
| *Escherichia coli* | 2.27 | 1.99 | 1.26 | 1.76 | 36.78 |
| *Blautia obeum* | 0.98 | 2.51 | 1.15 | 1.61 | 38.38 |
| *Ruminococcus gnavus* | 2.48 | 0.04 | 1.13 | 1.58 | 39.96 |
| *Bacteroides uniformis* | 0.14 | 2.55 | 1.11 | 1.55 | 41.51 |
| *Holdemanella biformis* | 2.01 | 0.45 | 1.09 | 1.52 | 43.03 |
| *Blautia faecis* | 2.12 | 2.30 | 0.98 | 1.38 | 44.40 |
| *Faecalibacterium duncaniae* | 2.09 | 2.15 | 0.98 | 1.37 | 45.78 |
| *Enterococcus 26* | 2.07 | 0.31 | 0.98 | 1.37 | 47.15 |
| *Faecalibacterium longum* | 0.42 | 2.19 | 0.96 | 1.35 | 48.49 |
| *Ruminococcus faecis* | 2.27 | 1.61 | 0.96 | 1.34 | 49.84 |
| *Faecalibacterium prausnitzii* | 0.00 | 2.04 | 0.95 | 1.32 | 51.16 |

Taxa identified as core are highlighted in orange, with satellite taxa highlighted in grey. The mean relative abundance (%) across both groups is given, alongside the percentage contribution which is the mean dissimilarity of taxa divided by the mean dissimilarity (71.26%) across samples. Cumulative percent does not equal 100% as the list is not exhaustive, rather the taxa that make up 50% of dissimilarity between groups. Given the length of the 16S gene regions sequenced, taxon identification should be considered putative.

To investigate the relationships between participant clinical variables and microbiota composition, redundancy analysis (RDA) was performed. Within comparisons between pwCF following extended ETI and the healthy control participants (Table 3), whole microbiota variability was explained by (in ascending order) the presence of CF disease, antibiotic usage, and age. For the core taxa analysis, CF disease was the primary driver of variation, with sex also contributing. CF disease was also the main contributor of variation in the satellite taxa, followed by antibiotic usage and patient small bowel water content (SBWC). When relating variability of the microbiota solely within pwCF (from baseline to extended ETI) (Table S10), multiple variables significantly explained the variation in the whole microbiota, including disease severity (FEV1%), BMI, antibiotic usage, sex, and SBWC. Excluding BMI, these variables also significantly explained variation across the core taxa. For the satellite taxa analysis, Age and ETI treatment length also significantly explained some of the variation. Species RDA biplots were then plotted to visualise how the taxa driving differences across pwCF and controls related to significant clinical variables identified from the RDA approach. Between the healthy controls and pwCF on extended ETI (Figure 3), a group of taxa containing *Faecalibacterium* and *Bacteroides* members collectively clustered strongly away from CF disease and the usage of antibiotics, whilst taxa such as *Blautia* sp. (OTU 1), *Ruminoccocus gnavus, Enterococcus* sp. (OTU 26), and *Eubacterium hallii* demonstrated the opposite trend. Some taxa were explained primarily by both participant age and sex. This included *Blautia luti*, *Dorea longicatena*, *Holdemanella biformis*, *Bacteroides dorei*, *Escherichia coli*, and *Bifidobacterium adolescentis*. In terms of SBWC, this explained the variation across a group of taxa including *Fusicatenibacter saccharivorans* and *Gemmiger formicilis*. Within pwCF exclusively (baseline to extended ETI), different taxa and variables comprised the RDA biplot due to outcomes from the previous SIMPER and RDA analyses (Figure S2). Some similar trends were observed however, including the strong association of *Enterococcus* sp. (OTU 24) with antibiotic usage and *Blautia* sp. (OTU 1) away from more mild CF disease.

**Figure 3.**
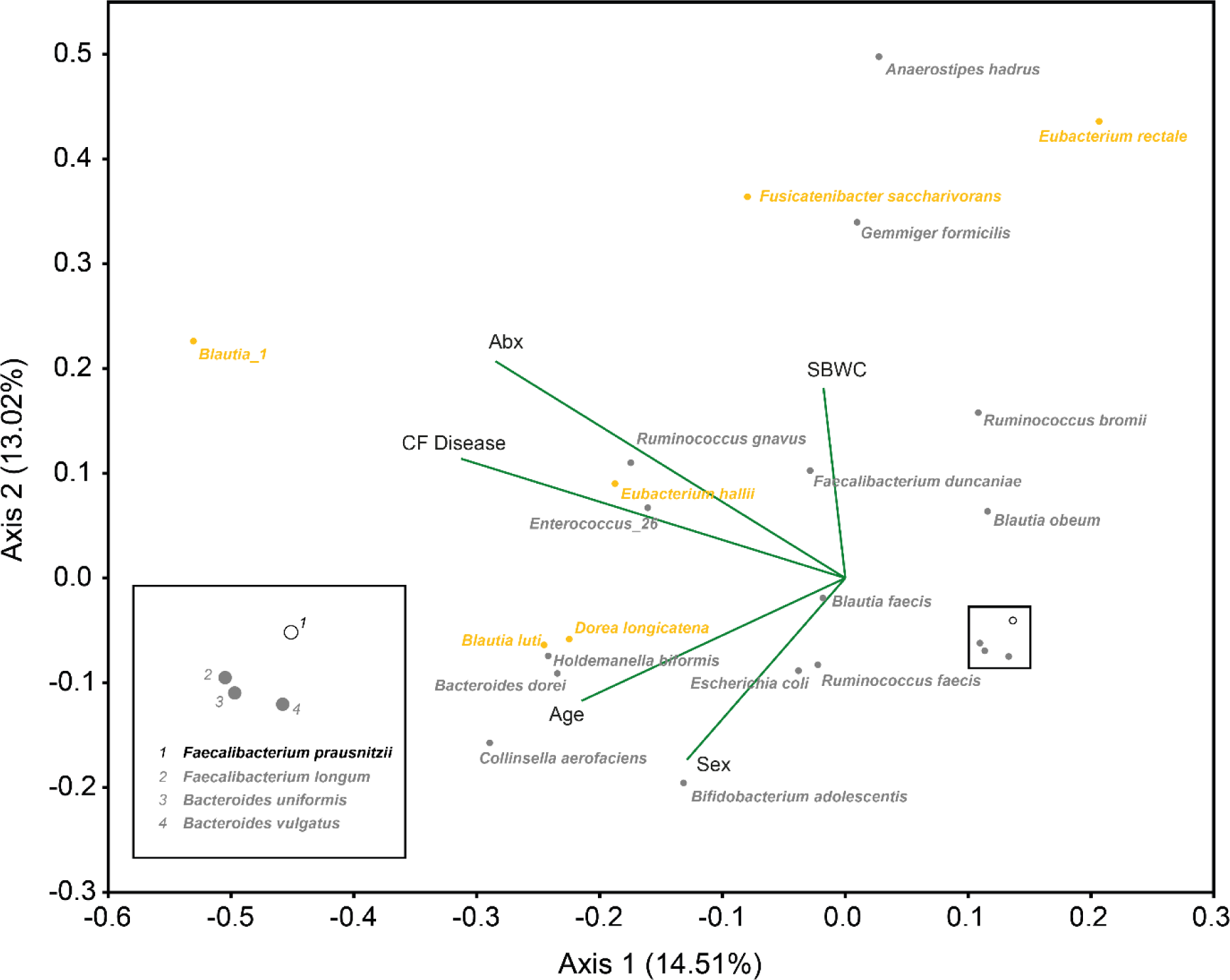
Redundancy analysis species biplots for the whole microbiota. The 24 taxa contributing most to the dissimilarity (cumulatively > 50%) between healthy control and pwCF samples following extended ETI therapy from the SIMPER analysis (Table 2) are shown independently of the total number of ASVs identified (531). Orange points represent taxa that were identified as core for the pwCF group following extended ETI therapy, grey points are satellite, and the white (black stroke) points represent taxa that were absent. Biplot lines depict clinical variables that significantly account for total variation in taxa relative abundance within whole microbiota analysis at the *p* ≤ 0.05 level (Table 3). Species plots depict the strength of explanation provided by the given clinical variables, with taxa shown in the same direction of a particular clinical variable considered to have a higher value than those that are not. ‘Abx’ – Antibiotics during sampling period, ‘SBWC’ – Small bowel water content corrected for body surface area. The percentage of microbiota variation explained by each axis is given in parentheses.

**Table 3.** Redundancy analysis to explain percent variation across whole microbiota, core, and satellite taxa of the significant clinical variables across healthy control participants and pwCF receiving 17+ months (extended) ETI therapy.

|  | Microbiota |  |  | Core taxa |  |  | Satellite taxa |  |  |
| --- | --- | --- | --- | --- | --- | --- | --- | --- | --- |
|  | Var.<br>Exp<br>(%) | pseudo-*F* | *P*<br>(adj) | Var.<br>Exp<br>(%) | pseudo-*F* | *P*<br>(adj) | Var.<br>Exp<br>(%) | pseudo-*F* | *P*<br>(adj) |
| Age | 10.1 | 2.0 | 0.008 |  |  |  |  |  |  |
| Antibiotics | 10.6 | 2.0 | 0.018 |  |  |  | 10.8 | 1.9 | 0.012 |
| CF Disease | 13.3 | 2.3 | 0.002 | 27.5 | 5.7 | 0.002 | 16.0 | 3.3 | 0.002 |
| SBWC |  |  |  |  |  |  | 10.8 | 1.8 | 0.050 |
| Sex |  |  |  | 10.0 | 2.2 | 0.028 |  |  |  |
| Total | **23.9** |  |  | **37.5** |  |  | **37.6** |  |  |

Both the composition and absolute quantification of SCFAs from pwCF and healthy control samples were also investigated (Figure S3). Combined, the relative abundance of acetic, propionic, and butyric acid accounted for the vast majority of SCFAs across both the pwCF ETI treatment periods (94.0% ± 4.5, Mean ± SD) and healthy controls (89.6% ± 4.8, Mean ± SD). There were no significant differences from baseline, or subsequent sampling periods, in the absolute concentration or relative abundance of SCFAs across pwCF receiving ETI therapy (Figure S3, Tables S11-12). There were, however, significant differences between ETI samples and healthy controls (Figure S3, Table S13-14). This included significant increases in both absolute concentration and relative abundance of valeric (Figure S3 A&C) and heptanoic acid (Figure S3 B&D) within healthy control samples compared with all ETI sampling periods (*P* < 0.05 in all instances), with hexanoic acid also significantly increased compared to pwCF, except those on extended ETI (*P* = 0.143) (Figure S3 B&D). On the contrary, the only significantly increased SCFA in pwCF was butyric acid, with significantly higher concentrations measured during the extended ETI sampling period as compared to health controls (*P* = 0.015) (Figure S3A).

The overall composition of SCFAs remained similar in pwCF throughout ETI therapy but remained significantly different from healthy controls during both the 6 months and extended ETI therapy periods (*P* < 0.05 in all instances) (Table S15). Over 50% of this dissimilarity was driven by acetic and butyric acid in both cases (Table S16). Exploratory RDA revealed that only a few SCFA, namely propionic, butyric, and valeric acid could significantly explain any variation across the microbiota composition across all samples, and to a minimal extent (Table S17). RDA biplots were constructed to visualise the relationships between SIMPER taxa and these SCFAs within both pwCF and healthy controls (Figure S4). Results were mixed, with high variability across particular genera, including *Bacteroides*, *Blautia*, *Ruminococcus,* whilst others such as *Faecalibacterium* and *Eubacterium* behaved similarly and were more closely clustered. Other taxa were strongly dissociated with the SCFAs from the RDA biplot model, including *Blautia* spp. (OTU 1) and *Fusicatenibacter saccharivorans*.

Finally, when comparing symptoms and gut function across pwCF and healthy controls, it was evident that ETI therapy did not significantly improve PAC-SYM scores irrespective of treatment length (*P* > 0.05 in all instances) (Table S18). In terms of functionality, no significant differences in oro-caecal transit time (OCTT) (*P* = 0.842) or SBWC (*P* = 0.064) between pwCF in this cohort undergoing microbiota analyses and healthy controls occurred, however the latter was markedly increased (Table S19).

## 4. Discussion

As the efficacy of CFTR modulators continues to increase in alleviating respiratory complications of CF, there is need to clarify whether patient improvements further translate at the site of the intestinal tract. This includes both GI abnormalities and patient symptoms following CFTR modulator treatment, including the effects upon the gut microbiota, for which data remain scarce and to the best of our knowledge has not been characterised in pwCF following ETI therapy. Here we examined the impact of ETI on the gut microbiota and their associated metabolites, utilising 16S rRNA sequencing and integrated targeted metabolomics to profile and quantify faecal levels of short-chain fatty acids. Overall, our results indicate that ETI therapy does impact upon microbiota structure in pwCF, but this remains distinctly different to that of healthy control participants as observed previously [8,32,33].

With respect to microbiota diversity, we demonstrate a significant shift in the core taxa of pwCF following extended ETI therapy, which translated into a positive trajectory of whole microbiota diversity by the end of the study period. Despite this, diversity was not significantly different following ETI, as shown previously with less efficacious CFTR modulator treatments encompassing shorter administration periods [15–17,19]. Interestingly, Kristensen et al. did observe significant increases in faecal microbiota diversity [18], but only following extended (12 month) Ivacaftor therapy in a group of pwCF harbouring the S1251N mutation. Unlike others [15–17,19], they also found that microbiota composition changed significantly over time, with samples obtained following extended Ivacaftor more tightly clustered [18]. In the current study, the core taxa and whole microbiota became more similar in pwCF as ETI progressed, also increasing in similarity to healthy controls. There remained, however, a significant difference in microbiota composition to controls as previously observed throughout life in pwCF [7–9,24].

The significant difference in microbiota composition maintained between pwCF and controls following extended ETI was driven by taxa previously associated with SCFA production in the gut, including members of the genera *Faecalibacterium*, *Eubacterium*, and *Blautia* [34,35]. Across pwCF specifically, *Faecalibacterium* remained a satellite taxa constituent and did not substantially contribute to any changes in the community from baseline, whilst relative abundances of *Eubacterium* were generally increased, and *Blautia* species fluctuated over the treatment period in pwCF. This raises the possibility of increased fitness and subsequent proliferation of particular taxa following altered intestinal fluidity subsequent to CFTR modulation, alongside decreased susceptibility to additional CF-associated perturbations that typically impacts the rarer satellite taxa. When incorporating targeted metabolomics to understand any functional changes relating to the aforementioned taxa, we identified no key differences in pwCF following ETI treatment, further extending with healthy controls for acetic, propionic, isobutyric, butyric and isovaleric acid. These findings are in agreement with Baldwin-Hunter et al., who recently analysed colonic aspirates from adult pwCF and controls undergoing surveillance colonoscopy [36]. Collectively, this may support elements of functional redundancy surrounding major SCFA production in the distal colon in pwCF [37], and is further supported by the low microbial variation explained here by faecal SCFA levels when both pwCF and control samples were collated. In contrast, Vernocchi et al. did demonstrate significant reductions in major SCFAs across children with CF as compared to controls [38], which raises the possibility of age-dependent functional redundancy. Similar to Baldwin-Hunter et al. again [36], both valeric and hexanoic acid concentration and relative abundances were significantly larger in healthy control participants compared to pwCF. Furthermore, valeric acid did significantly explain variation in the microbiota across all samples collated for analysis, with those associated taxa typically satellite members of the microbiota in pwCF. Any potential implications of this remain to be clarified in CF as valeric and hexanoic acid have previously been shown to significantly decrease within intestinal pro-inflammatory environments as compared to healthy controls [39].

Furthering our analyses to reveal any relationships between clinical data, gut function, and microbiota structure across participants, we observed a strong impact of CF disease, antibiotic exposure, and age upon microbiota composition similar to our previous work [24]. CF disease was the primary explanator of the core taxa, which is unsurprising given the strong associations between CFTR genotype alone and gut microbiota composition [11]. Alongside the whole microbiota, antibiotic usage was a principal explanator of satellite taxa composition across participants, again underpinning the relevance of this partitioned community towards wider microbiota organisation and structure. Within the exclusive analysis of pwCF, antibiotic usage was a prominent explanator of the bacterial community across all partitions, second only to age within the satellite taxa analysis and explaining more variance than CF disease severity and treatment length of ETI. Indeed, antibiotics have previously been shown to significantly impact community composition across pwCF also taking CFTR modulators, to which the latter had relatively little impact [17]. Given the speculated systemic impact of pulmonary antibiotics in-turn affecting the gut microbiota [8,24,38,40,41], it still remains in question whether this will persist following sustained CFTR modulator therapy that commences earlier in life across pwCF [42]. In this cohort, not all participants were undertaking antibiotic therapy, including those on extended ETI. Of the pwCF that were administered antibiotics however, levels of exposure were at least maintained during the extended ETI period. Gut function also continues to explain the microbiota variance across pwCF and controls [24]. SBWC was not significantly altered following ETI treatment and remains elevated across pwCF [23], significantly explaining the satellite taxa composition across all participants in this study. The physiological mechanisms behind this increased SBWC are proposed to result from gastro-ileal abnormalities in CF [43], including delayed ileal-emptying of small bowel content which might have consequences for downstream taxa in the large intestine.

Relating bacterial taxa from our SIMPER analysis with principal drivers of microbiota variation determined by the RDA approach revealed associations across species both typically identified as favourable and adverse to the host with CF disease and antibiotic usage. This included *Blautia* spp. and *Eubacterium hallii* which contain the functional capacity for anti-inflammatory butyrate synthesis from a range of substrates [44], but also *Enterococcus* spp. and *Ruminococcus gnavus*, which have both been shown to be increased in the CF gut and associate with intestinal inflammation [9,45,46], perhaps indicative of acquired resistance to recurrent antibiotic regimens in CF [47]. Within pwCF specifically, the relationships between disease severity (determined by FEV1%) and intestinal microbial composition may suggest the need to further understand the putative lung-gut axis in CF [6,48,49]. The implications of any taxa change upon host immunology at the site of the intestinal tract remains to be clarified and moving forward will be aided by the thorough integration of faecal and local intestinal inflammatory data. Across all participants, age and sex were also explanators of the whole microbiota and core taxa respectively, associating with taxa such as *Escherichia coli* and *Bifidobacterium adolescentis*. This may be explained in part by the ratio of males to females, alongside the wide age range of our participants, with the relative abundance of the aforementioned impacted by age across both CF and general life [40,50–52].

Alongside the microbiota analyses, it was evident that patient symptoms, as measured by overall PAC-SYM scores, were not improved upon treatment with ETI. This is consistent with our previous cohort undertaking Tezacaftor/Ivacaftor therapy [19], yet Mainz et al. have recently described improvements within pwCF GI symptoms upon ETI therapy when utilising the tailored CFAbd questionnaire [53]. Intestinal inflammation is a common occurrence in pwCF [54–56]. Unfortunately, we were unable to incorporate faecal calprotectin data into this study due to limited measurement across participants. Previous modulator studies have reported mixed results in the reduction of faecal calprotectin [16,57,58]. However, with ETI therapy specifically, there is evidence of a reduction following 6 months of administration [59].

Collectively, our microbiota findings indicate a positive impact of ETI therapy upon core taxa and whole microbiota structure in pwCF. The significant differences that persist between the CF and healthy control microbiota suggests changes are further required across the CF satellite taxa to bridge this gap, which make up a significant portion of the relative community abundance [24], and are typically related to random environmental perturbations or atypical habitats [60,61]. Based on this, the modulation of CFTR alone may not be sufficient to rescue a microbiota signature observed across controls within pwCF, whereby the modulation of such alternate clinical factors primarily explaining the satellite taxa composition may be also required for an enhanced response to CFTR modulator therapy. This encompasses pulmonary antibiotic usage, which was a key explanator across our multivariate analyses and of high prevalence across pwCF undertaking extended ETI. Though not investigated in the current study, additional research into the ability of specific dietary intake and probiotic usage to shape the microbiota and patient outcomes is desirable [62–64], especially since resilience of the gut microbiota has been recently suggested following diet and exercise intervention in pwCF [65]. It could be that long-term substantial changes to such factors are necessary for subsequent beneficial changes to the intestinal microbiome in an era of modulator therapy.

Comprehensively understanding the impact of more efficacious CFTR modulator therapy itself remains a top research priority in CF, including the identification of novel compounds or biomarkers to enhance our understanding of disease management [66,67]. The latter is likely to be aided by continued integration of additional omics techniques to fully elucidate not only the genetic potential, but measurable functional alterations upon CFTR modulator usage and changes to the CF lifestyle. This should include the impact of pulmonary antibiotic towards CF intestinal metabolomic and/or metaproteomic signatures, given their common, persistent use in clinic and at home. This may extend further to the utilisation of model experimental systems [68], which with tighter experimental control will aid our future understanding of the aforementioned. Aside from the clear caveat of group size in our pwCF cohort receiving extended ETI therapy, we acknowledge the relatively large sampling gap between this period and the preceding sampling point of 6 months within this pilot study. Nonetheless, our extended sampling period highlights the value of further understanding alternate factors to the sole presence of disease in altering the CF microbiota, patient symptoms and intestinal abnormalities, as treatment with CFTR modulators alone may not be optimal to achieve this goal in CF.

## 5. Conclusion

In conclusion, we have demonstrated that ETI therapy in pwCF does lead to change in microbiota characteristics towards that observed in healthy controls following extended administration, albeit with negligible difference upon SCFA levels and other clinical outcomes. Collectively the composition of the microbiota and functional metabolites remained significantly different from healthy controls following extended therapy. We posit that the trajectory of the CF gut microbiota towards that observed in the healthy controls was predominately impeded by antibiotics administered to combat respiratory infection. We recommend future work should employ a combination of frequent/prolonged patient cohort studies and experimental model system approaches to gain a deeper mechanistic understanding of the effects of both the CFTR modulator therapies and pulmonary antibiotics on the temporal dynamics of gut microbiota composition functioning. In turn this could then be related to alleviating and improving abnormalities of the intestinal tract and patient symptoms in CF.

## Supporting information

Marsh Supplementary MedRxiv

## Data Availability

All data produced in the present study are available upon reasonable request to the authors

## Author contributions

CvdG, DR, ARS, GM, and RM conceived the microbiome study. RM, and LH performed microbiota sample processing and analysis. RM and CDS carried out the metabolomic analysis. RM, DR, and CvdG performed the data and statistical analysis. AY, ND, CH, CN, GM, and AS were responsible for sample collection, clinical data collection, care records, and documentation. RM, AY, and CvdG verified the underlying data. RM, DR, and CvdG were responsible for the creation of the original draft of the manuscript. RM, GM, DR, ARS, and CvdG contributed to the development of the final manuscript. CvdG is the guarantor of this work. All authors read and approved the final manuscript.

## Acknowledgements

We would like to thank Neele Dellschaft, Caroline Hoad, Luca Marciani, and Penny Gowland of the Sir Peter Mansfield Imaging Centre, University of Nottingham, who acquired the original MRI data for healthy control participants [23].

## References

[1] S. Smith, N. Rowbotham, G. Davies, K. Gathercole, S.J. Collins, Z. Elliott, S. Herbert, L. Allen, C. Ng, A. Smyth, How can we Relieve Gastrointestinal Symptoms in people with Cystic Fibrosis? An International Qualitative Survey, BMJ Open Respir. Res. 7 (2020) e000614. 10.1136/bmjresp-2020-000614.

[2] M. Lopes-Pacheco, CFTR Modulators: The Changing Face of Cystic Fibrosis in the Era of Precision Medicine, Front. Pharmacol. 10 (2020) 1662. 10.3389/fphar.2019.01662.

[3] Cystic Fibrosis Trust, UK Cystic Fibrosis Trust, UK Cystic Fibrosis Registry. 2022 Annual Data Report, London, 2023.

[4] Cystic Fibrosis Refresh Top 10 priorities (priority setting in association with the JLA), (2022). https://www.jla.nihr.ac.uk/priority-setting-partnerships/cystic-fibrosis-refresh/top-10-priorities.htm.

[5] N.J. Rowbotham, S. Smith, Z.C. Elliott, B. Cupid, L.J. Allen, K. Cowan, L. Allen, A.R. Smyth, A Refresh of the Top 10 Research Priorities in Cystic Fibrosis, Thorax. 78 (2023) 840–843. 10.1136/thorax-2023-220100.

[6] J.C. Madan, D.C. Koestle, B.A. Stanton, L. Davidson, L.A. Moulton, M.L. Housman, J.H. Moore, M.F. Guill, H.G. Morrison, M.L. Sogin, T.H. Hampton, M.R. Karagas, P.E. Palumbo, J.A. Foster, P.L. Hibberd, G.A. O’Toole, Serial Analysis of the Gut and Respiratory Microbiome, MBio. 3 (2012) e00251–12. 10.1128/mBio.00251-12.Editor.

[7] M.J. Coffey, S. Nielsen, B. Wemheuer, N.O. Kaakoush, M. Garg, B. Needham, R. Pickford, A. Jaffe, T. Thomas, C.Y. Ooi, Gut Microbiota in Children With Cystic Fibrosis: A Taxonomic and Functional Dysbiosis, Sci. Rep. 9 (2019) 18593. 10.1038/s41598-019-55028-7.

[8] D.G. Burke, F. Fouhy, M.J. Harrison, M.C. Rea, P.D. Cotter, O.O. Sullivan, C. Stanton, C. Hill, F. Shanahan, B.J. Plant, R.P. Ross, The Altered Gut Microbiota in Adults with Cystic Fibrosis, BMC Microbiol. 17 (2017) 58. 10.1186/s12866-017-0968-8.

[9] M. Kristensen, S.M.P.J. Prevaes, G. Kalkman, G.A. Tramper-Stranders, R. Hasrat, K.M. de Winter-de Groot, H.M. Janssens, H.A. Tiddens, M. van Westreenen, E.A.M. Sanders, B. Arets, B. Keijser, C.K. van der Ent, D. Bogaert, Development of the Gut Microbiota in Early Life: The Impact of Cystic Fibrosis and Antibiotic Treatment, J. Cyst. Fibros. 19 (2020) 553–561. 10.1016/j.jcf.2020.04.007.

[10] D. Debray, H. El Mourabit, F. Merabtene, L. Brot, D. Ulveling, Y. Chrétien, D. Rainteau, I. Moszer, D. Wendum, H. Sokol, C. Housset, Diet-Induced Dysbiosis and Genetic Background Synergize With Cystic Fibrosis Transmembrane Conductance Regulator Deficiency to Promote Cholangiopathy in Mice, Hepatol. Commun. 2 (2018) 1533–1549. 10.1002/hep4.1266.

[11] S.M. Meeker, K.S. Mears, N. Sangwan, M.J. Brittnacher, E.J. Weiss, P.M. Treuting, N. Tolley, C.E. Pope, K.R. Hager, A.T. Vo, J. Paik, C.W. Frevert, H.S. Hayden, L.R. Hoffman, S.I. Miller, A.M. Hajjar, CFTR Dysregulation Drives Active Selection of the Gut Microbiome, PLoS Pathog. 16 (2020) e1008251. 10.1371/journal.ppat.1008251.

[12] M.B. De Freitas, E.A.M. Moreira, C. Tomio, Y.M.F. Moreno, F.P. Daltoe, E. Barbosa, N.L. Neto, V. Buccigrossi, A. Guarino, Altered Intestinal Microbiota Composition, Antibiotic Therapy and Intestinal Inflammation in Children and Adolescents with Cystic Fibrosis, PLoS One. 13 (2018) e0198457. 10.1371/journal.pone.0198457.

[13] T. Flass, S. Tong, D.N. Frank, B.D. Wagner, C.E. Robertson, C.V. Kotter, R.J. Sokol, E. Zemanick, F. Accurso, E.J. Hoffenberg, M.R. Narkewicz, Intestinal Lesions are Associated with Altered Intestinal Microbiome and are More Frequent in Children and Young Adults with Cystic Fibrosis and Cirrhosis, PLoS One. 10 (2015) e0116967. 10.1371/journal.pone.0116967.

[14] G. Dayama, S. Priya, D.E. Niccum, A. Khoruts, R. Blekhman, Interactions Between the Gut Microbiome and Host Gene Regulation in Cystic Fibrosis, Genome Med. 12 (2020) 12. 10.1186/s13073-020-0710-2.

[15] C.Y. Ooi, S.A. Syed, L. Rossi, M. Garg, B. Needham, J. Avolio, K. Young, M.G. Surette, T. Gonska, Impact of CFTR Modulation with Ivacaftor on Gut Microbiota and Intestinal Inflammation, Sci. Rep. 8 (2018). 10.1038/s41598-018-36364-6.

[16] N.J. Ronan, G.G. Einarsson, J. Deane, F. Fouhy, M. Rea, C. Hill, F. Shanahan, J.S. Elborn, R.P. Ross, M. McCarthy, D.M. Murphy, J.A. Eustace, T. Mm, C. Stanton, B.J. Plant, Modulation, Microbiota and Inflammation in the adult CF Gut: A Prospective Study, J. Cyst. Fibros. 21 (2022) 837–843. 10.1016/j.jcf.2022.06.002.

[17] C.E. Pope, A.T. Vo, H.S. Hayden, E.J. Weiss, S. Durfey, S. McNamara, A. Ratjen, B. Grogan, S. Carter, L. Nay, M.R. Parsek, P.K. Singh, E.F. McKone, M.L. Aitken, M.R. Rosenfeld, L.R. Hoffman, Changes in Fecal Microbiota with CFTR Modulator Therapy: A Pilot Study, J. Cyst. Fibros. 20 (2021) 742–746. 10.1016/j.jcf.2020.12.002.

[18] M.I. Kristensen, K.M. de Winter-De Groot, G. Berkers, M.L.J.N. Chu, K. Arp, S. Ghijsen, H.G.M. Heijerman, H.G.M. Arets, C.J. Majoor, H.M. Janssens, R. van der Meer, D. Bogaert, C.K. van der Ent, Individual and Group Response of Treatment with Ivacaftor on Airway and Gut Microbiota in People with CF and a s1251n Mutation, J. Pers. Med. 11 (2021) 350. 10.3390/jpm11050350.

[19] R. Marsh, C. Dos Santos, L. Hanson, C. Ng, G. Major, A.R. Smyth, D. Rivett, C. van der Gast, Tezacaftor/Ivacaftor Therapy has Negligible Effects on the Cystic Fibrosis Gut Microbiome., Microbiol. Spectr. 11 (2023) e0117523. 10.1128/spectrum.01175-23.

[20] C. Tetard, M. Mittaine, S. Bui, F. Beaufils, P. Maumus, M. Fayon, P.-R. Burgel, T. Lamireau, L. Delhaes, E. Mas, R. Enaud, Reduced Intestinal Inflammation With Lumacaftor/Ivacaftor in Adolescents With Cystic Fibrosis, J. Pediatr. Gastroenterol. Nutr. 71 (2020) 778–781.

[21] V.A. Stallings, N. Sainath, M. Oberle, C. Bertolaso, J.I. Schall, Energy Balance and Mechanisms of Weight Gain with Ivacaftor Treatment of Cystic Fibrosis Gating Mutations, J. Pediatr. 201 (2018) 229–237.e4. 10.1016/j.jpeds.2018.05.018.

[22] S.N. Dawood, A.M. Rabih, A. Niaj, A. Raman, M. Uprety, M.J. Calero, M.R.B. Villanueva, N. Joshaghani, N. Villa, O. Badla, R. Goit, S.E. Saddik, L. Mohammed, Newly Discovered Cutting-Edge Triple Combination Cystic Fibrosis Therapy: A Systematic Review., Cureus. 14 (2022) e29359. 10.7759/cureus.29359.

[23] C. Ng, N.S. Dellschaft, C.L. Hoad, L. Marciani, L. Ban, A.P. Prayle, H.L. Barr, A. Jaudszus, J.G. Mainz, R.C. Spiller, P. Gowland, G. Major, A.R. Smyth, Postprandial Changes in Gastrointestinal Function and Transit in Cystic Fibrosis Assessed by Magnetic Resonance Imaging, J. Cyst. Fibros. 20 (2021) 591–597. 10.1016/j.jcf.2020.06.004.

[24] R. Marsh, H. Gavillet, L. Hanson, C. Ng, M. Mitchell-Whyte, G. Major, A.R. Smyth, D. Rivett, C. van der Gast, Intestinal Function and Transit Associate with Gut Microbiota Dysbiosis in Cystic Fibrosis., J. Cyst. Fibros. 21 (2022) 506–513. 10.1016/j.jcf.2021.11.014.

[25] L. Frank, L. Kleinman, C. Farup, L. Taylor, P.J. Miner, Psychometric Validation of a Constipation Symptom Assessment Questionnaire., Scand. J. Gastroenterol. 34 (1999) 870– 877. 10.1080/003655299750025327.

[26] M.A. Gorzelak, S.K. Gill, N. Tasnim, Z. Ahmadi-Vand, M. Jay, D.L. Gibson, Methods for Improving Human Gut Microbiome Data by Reducing Variability through Sample Processing and Storage of Stool, PLoS One. 10 (2015) e0134802. 10.1371/journal.pone.0134802.

[27] G.B. Rogers, L. Cuthbertson, L.R. Hoffman, P.A.C. Wing, C. Pope, D.A.P. Hooftman, A.K. Lilley, A. Oliver, M.P. Carroll, K.D. Bruce, C.J. Van Der Gast, Reducing Bias in Bacterial Community Analysis of Lower Respiratory Infections, ISME J. 7 (2013) 697–706. 10.1038/ismej.2012.145.

[28] B.J. Callahan, P.J. McMurdie, M.J. Rosen, A.W. Han, A.J.A. Johnson, S.P. Holmes, DADA2: High Resolution Sample Inference from Illumina Amplicon Data, Nat. Methods. 13 (2016) 4–5. 10.1038/nmeth.3869.DADA2.

[29] N.M. Davis, D.M. Proctor, S.P. Holmes, D.A. Relman, B.J. Callahan, Simple Statistical Identification and Removal of Contaminant Sequences in Marker-Gene and Metagenomics Data, Microbiome. 6 (2018) 226. 10.1186/s40168-018-0605-2.

[30] Ø. Hammer, D. A.T. Harper, P.D. Ryan, PAST: Paleontological Statistics Software Package for Education and Data Analysis, Palaeontol. Electron. 4 (2001) 9.

[31] C. ter Braak, P. Smilauer, CANOCO Reference Manual and User’s Guide: Software for Ordination., Ithaca: Microcomputer Power, 2012.

[32] F. Miragoli, S. Federici, S. Ferrari, A. Minuti, A. Rebecchi, E. Bruzzese, V. Buccigrossi, A. Guarino, M.L. Callegari, Impact of Cystic Fibrosis Disease on Archaea and Bacteria Composition of Gut Microbiota, FEMS Microbiol. Ecol. 93 (2017) fiw230. 10.1093/femsec/fiw230.

[33] O. Manor, R. Levy, C.E. Pope, H.S. Hayden, M.J. Brittnacher, R. Carr, M.C. Radey, K.R. Hager, S.L. Heltshe, B.W. Ramsey, S.I. Miller, L.R. Hoffman, E. Borenstein, Metagenomic Evidence for Taxonomic Dysbiosis and Functional Imbalance in the Gastrointestinal Tracts of Children with Cystic Fibrosis, Sci. Rep. 6 (2016) 22493. 10.1038/srep22493.

[34] M. Sakamoto, N. Sakurai, H. Tanno, T. Iino, M. Ohkuma, A. Endo, Genome-Based, Phenotypic and Chemotaxonomic Classification of Faecalibacterium Strains: Proposal of Three Novel Species Faecalibacterium duncaniae sp. nov., Faecalibacterium hattorii sp. nov. and Faecalibacterium gallinarum sp. nov., Int. J. Syst. Evol. Microbiol. 72 (2022) 005379. 10.1099/ijsem.0.005379.

[35] D. Parada Venegas, M.K. De la Fuente, G. Landskron, M.J. González, R. Quera, G. Dijkstra, H.J.M. Harmsen, K.N. Faber, M.A. Hermoso, Short Chain Fatty Acids (SCFAs)-Mediated Gut Epithelial and Immune Regulation and Its Relevance for Inflammatory Bowel Diseases, Front. Immunol. 10 (2019) 277. https://www.frontiersin.org/articles/10.3389/fimmu.2019.00277.

[36] B.L. Baldwin-Hunter, F.D. Rozenberg, M.K. Annavajhala, H. Park, E.A. DiMango, C.L. Keating, A.-C. Uhlemann, J.A. Abrams, The Gut Microbiome, Short Chain Fatty Acids, and Related Metabolites in Cystic Fibrosis Patients with and without Colonic Adenomas., J. Cyst. Fibros. 22 (2023) 738–744. 10.1016/j.jcf.2023.01.013.

[37] Y. Wang, L.E.X. Leong, R.L. Keating, T. Kanno, G.C.J. Abell, F.M. Mobegi, J.M. Choo, S.L. Wesselingh, A.J. Mason, L.D. Burr, G.B. Rogers, Opportunistic Bacteria Confer the Ability toFerment Prebiotic Starch in the Adult Cystic Fibrosis Gut, Gut Microbes. 10 (2019) 367–381. 10.1080/19490976.2018.1534512.

[38] P. Vernocchi, F. Del Chierico, A. Russo, F. Majo, M. Rossitto, M. Valerio, L. Casadei, A. La Storia, F. De Filippis, C. Rizzo, C. Manetti, P. Paci, D. Ercolini, F. Marini, E.V. Fiscarelli, B. Dallapiccola, V. Lucidi, A. Miccheli, L. Putignani, Gut microbiota signatures in cystic fibrosis: Loss of host CFTR function drives the microbiota enterophenotype, PLoS One. 13 (2018) e0208171. 10.1371/journal.pone.0208171.

[39] V. De Preter, K. Machiels, M. Joossens, I. Arijs, C. Matthys, S. Vermeire, P. Rutgeerts, K. Verbeke, Faecal Metabolite Profiling Identifies Medium-Chain Fatty Acids as Discriminating Compounds in IBD., Gut. 64 (2015) 447–458. 10.1136/gutjnl-2013-306423.

[40] G. Duytschaever, G. Huys, M. Bekaert, L. Boulanger, K. De Boeck, P. Vandamme, Dysbiosis of Bifidobacteria and Clostridium Cluster XIVa in the Cystic Fibrosis Fecal Microbiota, J. Cyst. Fibros. 12 (2013) 206–215. 10.1016/j.jcf.2012.10.003.

[41] R. Enaud, K.B. Hooks, A. Barre, T. Barnetche, C. Hubert, M. Massot, T. Bazin, H. Clouzeau, S. Bui, M. Fayon, P. Berger, P. Lehours, C. Bébéar, M. Nikolski, T. Lamireau, L. Delhaes, T. Schaeverbeke, Intestinal Inflammation in Children with Cystic Fibrosis Is Associated with Crohn’s-Like Microbiota Disturbances, J. Clin. Med. 8 (2019) 645. 10.3390/jcm8050645.

[42] R.H. Keogh, R. Cosgriff, E.R. Andrinopoulou, K.G. Brownlee, S.B. Carr, K. Diaz-Ordaz, E. Granger, N.P. Jewell, A. Lewin, C. Leyrat, D.K. Schlüter, M. van Smeden, R.D. Szczesniak, G.J. Connett, Projecting the Impact of Triple CFTR Modulator Therapy on Intravenous Antibiotic Requirements in Cystic Fibrosis using Patient Registry Data Combined with Treatment Effects from Randomised Trials, Thorax. 77 (2022) 873–881. 10.1136/thoraxjnl-2020-216265.

[43] N. Dellschaft, C. Hoad, L. Marciani, P. Gowland, R. Spiller, Small Bowel Water Content Assessed by MRI in Health and Disease: A Collation of Single-Centre Studies., Aliment. Pharmacol. Ther. 55 (2022) 327–338. 10.1111/apt.16673.

[44] A. Rivière, M. Selak, D. Lantin, F. Leroy, L. De Vuyst, Bifidobacteria and Butyrate-Producing Colon Bacteria: Importance and Strategies for their Stimulation in the Human Gut, Front. Microbiol. 7 (2016) 979. 10.3389/fmicb.2016.00979.

[45] M.T. Henke, D.J. Kenny, C.D. Cassilly, H. Vlamakis, R.J. Xavier, J. Clardy, Ruminococcus gnavus, a Member of the Human Gut Microbiome Associated with Crohn’s Disease, Produces an Inflammatory Polysaccharide, Proc. Natl. Acad. Sci. U. S. A. 116 (2019) 12672–12677. 10.1073/pnas.1904099116.

[46] F. Fouhy, N.J. Ronan, O. O’Sullivan, Y. McCarthy, A.M. Walsh, D.M. Murphy, M. Daly, E.T. Flanagan, C. Fleming, M. McCarthy, C. Shortt, J.A. Eustace, F. Shanahan, M.C. Rea, R.P. Ross, C. Stanton, B.J. Plant, A Pilot Study Demonstrating the Altered Gut Microbiota Functionality in Stable Adults with Cystic Fibrosis, Sci. Rep. 7 (2017) 6685. 10.1038/s41598-017-06880-y.

[47] S.L. Taylor, L.E.X. Leong, S.K. Sims, R.L. Keating, L.E. Papanicolas, A. Richard, F.M. Mobegi, S. Wesselingh, L.D. Burr, G.B. Rogers, The Cystic Fibrosis Gut as a Potential Source of Multidrug Resistant Pathogens., J. Cyst. Fibros. 20 (2021) 413–420. 10.1016/j.jcf.2020.11.009.

[48] I. Testa, O. Crescenzi, S. Esposito, Gut Dysbiosis in Children with Cystic Fibrosis: Development, Features and the Role of Gut-Lung Axis on Disease Progression., Microorganisms. 11 (2022) 9. 10.3390/microorganisms11010009.

[49] A.G. Hoen, J. Lia, L.A. Moulton, G.A. O’Toole, M.L. Housman, D.C. Koestler, M.F. Guill, J.H. Moore, P.L. Hibberd, H.G. Morrison, M.L. Sogin, M.R. Karagas, J.C. Madan, Associations Between Gut Microbial Colonization in Early Life and Respiratory Outcomes in Cystic Fibrosis, J. Pediatr. 167 (2015) 138–147.e1–3. 110.1016/j.bbi.2017.04.008.

[50] L.R. Hoffman, C.E. Pope, H.S. Hayden, S. Heltshe, R. Levy, S. McNamara, M.A. Jacobs, L. Rohmer, M. Radey, B.W. Ramsey, M.J. Brittnacher, E. Borenstein, S.I. Miller, Escherichia coli Dysbiosis Correlates with Gastrointestinal Dysfunction in Children with Cystic Fibrosis, Clin. Infect. Dis. 58 (2014) 396–399. 10.1093/cid/cit715.

[51] S. Matamouros, H.S. Hayden, K.R. Hager, M.J. Brittnacher, K. Lachance, E.J. Weiss, C.E. Pope, A.F. Imhaus, C.P. McNally, E. Borenstein, L.R. Hoffman, S.I. Miller, Adaptation of Commensal Proliferating Escherichia coli to the Intestinal Tract of Young Children with Cystic Fibrosis, Proc. Natl. Acad. Sci. U. S. A. 115 (2018) 1605–1610. 10.1073/pnas.1714373115.

[52] S. Arboleya, C. Watkins, C. Stanton, R.P. Ross, Gut Bifidobacteria Populations in Human Health and Aging., Front. Microbiol. 7 (2016) 1204. 10.3389/fmicb.2016.01204.

[53] J.G. Mainz, C. Zagoya, L. Polte, L. Naehrlich, L. Sasse, O. Eickmeier, C. Smaczny, A. Barucha, L. Bechinger, F. Duckstein, L. Kurzidim, P. Eschenhagen, L. Caley, D. Peckham, C. Schwarz, Elexacaftor-Tezacaftor-Ivacaftor Treatment Reduces Abdominal Symptoms in Cystic Fibrosis-Early results Obtained With the CF-Specific CFAbd-Score, Front. Pharmacol. 14 (2022) 1207356. 10.3389/fphar.2022.877118.

[54] A. Munck, Cystic Fibrosis: Evidence for Gut Inflammation., Int. J. Biochem. Cell Biol. 52 (2014) 180–183. 10.1016/j.biocel.2014.02.005.

[55] F. Beaufils, E. Mas, M. Mittaine, M. Addra, M. Fayon, L. Delhaes, H. Clouzeau, F. Galode, T. Lamireau, Increased Fecal Calprotectin Is Associated with Worse Gastrointestinal Symptoms and Quality of Life Scores in Children with Cystic Fibrosis, J. Clin. Med. 9 (2020) 4080.

[56] J. Dorsey, T. Gonska, Bacterial Overgrowth, Dysbiosis, Inflammation, and Dysmotility in the Cystic Fibrosis Intestine, J. Cyst. Fibros. 16 (2017) S14–S23. 10.1016/j.jcf.2017.07.014.

[57] V.A. Stallings, N. Sainath, M. Oberle, C. Bertolaso, J.I. Schall, Energy Balance and Mechanisms of Weight Gain with Ivacaftor Treatment of Cystic Fibrosis Gating Mutations, J. Pediatr. 201 (2018) 229–237.e4. 10.1016/j.jpeds.2018.05.018.

[58] C. Tétard, M. Mittaine, S. Bui, M. Beaufils, F., Maumus, P. Fayon, P.R. Burgel, T. Lamireau, L. Delhaes, E. Mas, R. Enaud, Reduced Intestinal Inflammation with Lumacaftor/Ivacaftor in Adolescents with Cystic Fibrosis, J. Pediatr. Gastroenterol. Nutr. 71 (2020) 778–781.

[59] S.J. Schwarzenberg, P.T. Vu, M. Skalland, L.R. Hoffman, C. Pope, D. Gelfond, M.R. Narkewicz, D.P. Nichols, S.L. Heltshe, S.H. Donaldson, C.A. Frederick, A. Kelly, J.E. Pittman, F. Ratjen, M. Rosenfeld, S.D. Sagel, G.M. Solomon, M.S. Stalvey, J.P. Clancy, S.M. Rowe, S.D. Freedman, Elexacaftor/Tezacaftor/Ivacaftor and Gastrointestinal Outcomes in Cystic Fibrosis: Report of Promise-GI., J. Cyst. Fibros. 22 (2022) 282–289. 10.1016/j.jcf.2022.10.003.

[60] C.J. van der Gast, A.W. Walker, F.A. Stressmann, G.B. Rogers, P. Scott, T.W. Daniels, M.P. Carroll, J. Parkhill, K.D. Bruce, Partitioning core and satellite taxa from within cystic fibrosis lung bacterial communities., ISME J. 5 (2011) 780–791. 10.1038/ismej.2010.175.

[61] A.E. Magurran, Species Abundance Distributions Over Time, Ecol. Lett. 10 (2007) 347–354. 10.1111/j.1461-0248.2007.01024.x.

[62] L.R. Caley, H. White, M.C. de Goffau, R.A. Floto, J. Parkhill, B. Marsland, D.G. Peckham, Cystic Fibrosis-Related Gut Dysbiosis: A Systematic Review., Dig. Dis. Sci. 68 (2023) 1797– 1814. 10.1007/s10620-022-07812-1.

[63] S. Esposito, I. Testa, E. Mariotti Zani, D. Cunico, L. Torelli, R. Grandinetti, V. Fainardi, G. Pisi, N. Principi, Probiotics Administration in Cystic Fibrosis: What Is the Evidence?, Nutrients. 14 (2022) 3160. 10.3390/nu14153160.

[64] I. McKay, J. van Dorst, T. Katz, M. Doumit, B. Prentice, L. Owens, Y. Belessis, S. Chuang, A. Jaffe, T. Thomas, M. Coffey, C.Y. Ooi, Diet and the gut-lung axis in cystic fibrosis - direct & indirect links., Gut Microbes. 15 (2023) 2156254. 10.1080/19490976.2022.2156254.

[65] R.L. Knoll, V.H. Jarquín-Díaz, J. Klopp, A. Kemper, K. Hilbert, B. Hillen, D. Pfirrmann, P. Simon, V. Bähner, O. Nitsche, S. Gehring, L. Markó, S.K. Forslund, K. Poplawska, Resilience and Stability of the CF-Intestinal and Respiratory Microbiome during Nutritional and Exercise Intervention, BMC Microbiol. 23 (2023) 44. 10.1186/s12866-023-02788-y.

[66] J.L. Taylor-Cousar, P.D. Robinson, M. Shteinberg, D.G. Downey, CFTR modulator therapy: transforming the landscape of clinical care in cystic fibrosis., Lancet. 402 (2023) 1171–1184. 10.1016/S0140-6736(23)01609-4.

[67] K.B. Hisert, S.E. Birket, J.P. Clancy, D.G. Downey, J.F. Engelhardt, I. Fajac, R.D. Gray, M.E. Lachowicz-Scroggins, N. Mayer-Hamblett, P. Thibodeau, K.L. Tuggle, C.E. Wainwright, K. De Boeck, Understanding and addressing the needs of people with cystic fibrosis in the era of CFTR modulator therapy, Lancet Respir. Med. 11 (2023) 916–931. 10.1016/S2213-2600(23)00324-7.

[68] G.A. O’Toole, A. Crabbé, R. Kümmerli, J.J. LiPuma, J.M. Bomberger, J.C. Davies, D. Limoli, V. V Phelan, J.B. Bliska, W.H. DePas, L.E. Dietrich, T.H. Hampton, R. Hunter, C.M. Khursigara, A. Price-Whelan, A. Ashare, R.A. Cramer, J.B. Goldberg, F. Harrison, D.A. Hogan, M.A. Henson, D.R. Madden, J.R. Mayers, C. Nadell, D. Newman, A. Prince, D.W. Rivett, J.D. Schwartzman, D. Schultz, D.C. Sheppard, A.R. Smyth, M.A. Spero, B.A. Stanton, P.E. Turner, C. van der Gast, F.J. Whelan, R. Whitaker, K. Whiteson, Model Systems to Study the Chronic, Polymicrobial Infections in Cystic Fibrosis: Current Approaches and Exploring Future Directions, MBio. 12 (2021) e0176321. 10.1128/mbio.01763-21.

