## Supplementary material for "Impact of extended Elexacaftor/Tezacaftor/Ivacaftor therapy on the gut microbiome in cystic fibrosis": Marsh Supplementary MedRxiv

#### Supplementary Methods

##### *Study participants and design*

Twenty-four pwCF, possessing either F508del homozygosity or compound heterozygotes with at least one copy of F508del, were initially recruited from Nottingham University Hospitals NHS Trust. Participants were asked to provide stool samples whilst attending the Sir Peter Mansfield Imaging Centre at the University of Nottingham. Ultimately, 20 pwCF (mean age at baseline, 21.0 ± 8.6 years, median age, 18 years provided samples available for inclusion in our analyses. Participants were asked to fast overnight and prior to the refraining from the use of laxatives and anti-diarrhoeals during their visit. Regular pancreatic enzyme replacement therapy and physiotherapy procedures were admitted.

Clinical assessments and sample collections were performed at baseline, with further samples collected following ETI therapy for 3 and 6 months, with some participants further providing samples over an additional 19.8 ± 2.0 months (mean ± SD) period (minimum 17+ months) to form this observational study. Whilst at clinic, gut physiology assessment utilising magnetic resonance imaging (MRI) was performed. This included measurement of oro-caecal transit time (OCTT) and small bowel water content (SBWC) following the consumption of a standardised meal. During attendance, participants also provided faecal samples, and completed the validated PAC-SYM to assess their gut symptoms [1].

From the recruited cohort, sampling faecal sampling was achieved for 19 pwCF 3 months post-ETI, 18 pwCF 6 months post-ETI, and finally in 7 pwCF for the extended (17+ months) ETI period. Faecal samples from 10 age-matched healthy controls (mean age, 21.4 ± 7.40 years, median age, 20 years) from our previous study were available for microbiota and metabolomic comparison also [2]. Participant numbers were randomised to maintain anonymity. A data file matching the randomly assigned study number to the participant identifiable information is held securely and password protected on a University of Nottingham cloud-based web server. Further clinical trial details can be found at clinicaltrials.gov, under the trial number NCT04618185 (https://clinicaltrials.gov/ct2/show/NCT04618185).

*PMA treatment prior to DNA extraction*

1 mg PMA (Biotium, CA, USA) was hydrated in 98 µl 20% dimethyl sulfoxide (DMSO) to give a working stock concentration of 20 mM. 300 mg of thawed stool was homogenised in 1.5 ml PBS, and then centrifuged at 3200 *x* g, for 5 minutes. The pellet was then resuspended in 0.5 ml PBS prior to subsequent PMA treatment. 1.25 µl of PMA (20 mM) was added to give a final concentration of 50 µM. Following the addition of PMA to samples in opaque Eppendorf tubes, PMA was mixed by vortexing for 10 seconds, followed by incubation for 15 minutes at room temperature (~20 °C). This step was repeated before the transfer of samples to clear 1.5 ml Eppendorf tubes and placement within a LED lightbox. Treatment occurred for 15 minutes to allow PMA intercalation into DNA from compromised bacterial cells. Samples were then centrifuged at 10,000 x g for 5 minutes. The supernatant was discarded, and the cellular pellet was resuspended in 200 µl PBS.

*Targeted amplicon sequencing – Bacterial 16S rRNA*

Step 1 amplicon generation with primers based on the universal primer sequences 515F & 926R as described by Walters et al. [3], was performed under the following conditions: Initial denaturation of 180 seconds at 98 ºC, followed by: 25 cycles of 30 seconds at 95 ºC, 30 seconds at 55 ºC and 30 seconds at 72 ºC. A final extension of 5 minutes at 72 ºC was also included to complete the reaction. Step 2, the addition of dual barcodes and Illumina adaptor sequences was performed under the following conditions: Initial denaturation of 30 seconds at 98 ºC, followed by: 10 cycles of 10 seconds at 98 ºC, 20 seconds at 62 ºC and 30 seconds at 72 ºC. A final extension of 2 minutes at 72 ºC was also included to complete this reaction. This resulted in the generation of an ~ 550 bp amplicon spanning the V4-V5 hypervariable regions of the 16S rRNA gene.

*Sequencing Controls and Library Pooling*

PCR & DNA extraction negative controls were implemented, alongside the use of mock community positive controls, which included a Gut Microbiome Standard (ZYMO RESEARCH™). Following Barcode attachment in the second PCR step, clean-up and subsequent amplicon size selection was performed with AMPure XP beads (Beckman Coulter™) and quantified using a Qubit™ dsDNA HS kit. Sample concentrations were then manually normalised, pooled, and diluted to the final library concentrations requited for use on the Illumina MiSeq system.

*Sequence processing and analysis*

Initial sequencing files were processed using cutadapt (Version 3.5.0) to trim and remove upstream adaptor sequences prior to sequence analysis [4]. DADA2 was used to demultiplex and remove primer sequences, validate the quality profiles of forward and reverse reads and subsequently trim, infer sequence variants, merge denoised paired-reads, remove chimeras, and finally assign taxonomy via Naive Bayesian Classifier implementation. This included the use of a dedicated human intestinal 16S rRNA reference database [5]. Unidentifiable ASVs were run through BLAST [6] (https://blast.ncbi.nlm.nih.gov/Blast.cgi) and matched appropriately based on query coverage where possible. Taxa with 2 ≥ reads for a single sample were removed and excluded from subsequent statistical analysis. ASVs from the same bacterial taxon were collapsed to form a single OTU for a given taxon. The R package decontam was used to remove any potential source of contamination across samples [7], utilising the prevalence-based contamination identification approach with a threshold classification of $P = 0.1$.

*GC-MS: Sample processing and SCFA preparation*

Faecal samples stored at -80ºC were ground in liquid nitrogen before 50 mg of ground faeces was added to 500 µl MS-grade water. Samples were lysed and homogenised utilising inert ZR BashingBead Lysis Tubes (Cambridge bioscience, UK), using the FastPrep-24 5G instrument (MP Biomedicals, California, USA) with two cycles at a speed of 6.0 m/s for 40 seconds each. Samples were then incubated at 4 °C whilst mixing for 30 minutes using an ELMI Intelli-Mixer™ RM-2L at 80 rpm. Samples were centrifuged at 13,000 x g for 30 mins. The supernatant containing faecal SCFAs was removed. 150 µl of supernatant was protonated with 5M HCl before the addition of anhydrous diethyl ether (1:1 v/v), samples were vortexed for 10 seconds, and incubated on ice for 5 mins. Following incubation, samples were mixed with the ELMI Intelli-Mixer™ RM-2L as before for 15 minutes, then centrifuged at 10,000 x g for 5 mins. The DE layer containing faecal SCFAs was transferred to a new Eppendorf tube pre-loaded with 25 mg Na2SO4. The remaining layer was then re-extracted with another 150 µl DE as before. Samples were equally pooled and then 40 ul was then transferred to a GC-MS vial, with the addition of 2 ul N, O-bis(trimethylsilyl) trifluoroacetamide (BSTFA). Samples were vortexed then incubated for 3 hours at 37 °C before loading onto the GC-MS. MS grade water processed in parallel was used as a blank sample to correct the background.

*GC-MS Analysis*

GC-MS analysis was carried out using an Agilent 7890B/5977 Single Quadrupole Mass Selective Detector (MSD) (Agilent Technologies) equipped with a non-polar HP-5ms Ultra Inert capillary column (30 m × 0.25 mm × 0.25 µm) (Agilent Technologies). The Agilent 7693 Autosampler was used to inject 1.0 µl of the derivatised sample in triplicate at a split ratio of 20:1 at 265 °C, with a solvent delay of 2 minutes 30 seconds. The initial oven temperature was held at 40 °C for 2 minutes, followed by a 10 °C/min temperature ramp to 140 °C, then increased to 300 °C at the rate of 40 °C/min and kept at this temperature for 6 minutes. Electron impact (EI) mode ionisation was utilised at 70 eV, with the instrumental parameters set at 230, 150 and 300 °C for source, quadrupole and interface temperatures, respectively. Selected ion monitoring (SIM) mode was used for quantification; all confirmation and target ions lists are summarised in Supplementary Table S2. Agilent MassHunter workstation version B.07.00 programs were used to perform post-run analyses. A $^{13}C$-short chain fatty acids stool mixture (Merck Life Science, Poole, UK) was used as the internal standard to normalise all spectra obtained prior to analyses. A $^{13}C$-short chain fatty acids stool mixture (Merck Life Science, Poole, UK) was used as the internal standard to normalise all spectra obtained prior to analyses. A volatile fatty acid mixture (Merck Life Science, UK) was used to construct calibration curves for the quantification of target metabolites.

*References*

[1] L. Frank, L. Kleinman, C. Farup, L. Taylor, P.J. Miner, Psychometric Validation of a Constipation Symptom Assessment Questionnaire., Scand. J. Gastroenterol. 34 (1999) 870–877. https://doi.org/10.1080/003655299750025327.

[2] R. Marsh, H. Gavillet, L. Hanson, C. Ng, M. Mitchell-Whyte, G. Major, A.R. Smyth, D. Rivett, C. van der Gast, Intestinal Function and Transit Associate with Gut Microbiota Dysbiosis in Cystic Fibrosis., J. Cyst. Fibros. 21 (2022) 506–513. https://doi.org/https://doi.org/10.1016/j.jcf.2021.11.014.

[3] W. Walters, E.R. Hyde, D. Berg-lyons, G. Ackermann, Improved Bacterial 16S rRNA Gene (V4 and V4-5) and Fungal Internal Transcribed Spacer Marker Gene Primers for Microbial Community Surveys, MSystems. 1 (2016) e00009-15. https://doi.org/10.1128/mSystems.00009-15.Editor.

[4] M. Martin, Cutadapt Removes Adapter Sequences from High-Throughput Sequencing Reads, EMBnet. J. 17 (2011) 10–12.

[5] J. Ritari, J. Salojärvi, L. Lahti, W.M. de Vos, Improved Taxonomic Assignment of Human Intestinal 16S rRNA Sequences by a Dedicated Reference Database, BMC Genomics. 16 (2015) 1056. https://doi.org/10.1186/s12864-015-2265-y.

[6] S.F. Altschul, T.L. Madden, A.A. Schäffer, J. Zhang, Z. Zhang, W. Miller, D.J. Lipman, Gapped BLAST and PSI-BLAST: A New Generation of Protein Database Search Programs., Nucleic Acids Res. 25 (1997) 3389–3402. https://doi.org/10.1093/nar/25.17.3389.

[7] N.M. Davis, D.M. Proctor, S.P. Holmes, D.A. Relman, B.J. Callahan, Simple Statistical

Identification and Removal of Contaminant Sequences in Marker-Gene and Metagenomics Data, Microbiome. 6 (2018) 226. https://doi.org/10.1186/s40168-018-0605-2.

### Supplementary Results

**Table 1**. Clinical metadata for pwCF and healthy control participants.

| | | | Genotype | | | | | | | | | Antibiotics | | | | | | |
|---|---|---|---|---|---|---|---|---|---|---|---|---|---|---|---|---|---|---|
| ID | Group | Treatment length (Months) | 1 | 2 | Sex | Age (Years) | BMI | PI | CFRD | FCP | FEV1% | Ami | Ant | Mac | Mon | Pol | Sul | Tet |
| 023 | pwCF | B | F508del | 1154insTC, p.Phe342HishX28 | M | 12-17 | 17.5 | Y | N | - | 94 | - | - | - | - | - | - | - |
| | | 3 | | | M | 12-17 | 18.0 | | | - | 89 | - | - | - | - | - | - | - |
| | | 6 | | | M | 12-17 | 18.4 | | | - | 94 | - | - | - | - | - | - | - |
| | | 20 | | | M | 12-17 | 19.0 | | | - | 89 | - | - | - | - | - | - | - |
| 046 | pwCF | B | F508del | F508del | F | 18-23 | 21.9 | Y | N | 14.1 | 57 | - | - | + | - | - | - | - |
| | | 3* | | | F | 18-23 | 21.2 | | | 7.0 | 71.4 | - | - | + | - | - | - | - |
| | | 6 | | | F | 18-23 | 21.4 | | | 7.0 | 73.5 | - | - | + | - | - | - | - |
| 194 | pwCF | B | F508del | F508del | M | 18-23 | 22.8 | Y | Y | <5 | 91 | - | - | - | - | - | - | - |
| | | 3 | | | M | 18-23 | 23.3 | | | <5 | 98.9 | - | - | - | - | - | - | - |
| | | 6 | | | M | 18-23 | 24.3 | | | <5 | 93.1 | - | - | - | - | - | - | - |
| 247 | pwCF | B | F508del | F508del | M | 12-17 | 20.8 | Y | N | 27.6 | 101 | - | - | - | - | - | - | - |
| | | 3 | | | M | 18-23 | 21.7 | | | - | - | - | - | - | - | - | - | - |
| 253 | pwCF | B | F508del | F508del | M | 12-17 | 21.8 | Y | N | 9.8 | 81 | - | - | + | - | - | - | - |
| | | 3 | | | M | 12-17 | 27.0 | | | - | 101.5 | - | - | + | - | - | - | - |
| | | 6 | | | M | 12-17 | 28.0 | | | - | 98.2 | - | - | + | - | - | - | - |
| 257 | pwCF | B | F508del | F508del | M | 12-17 | 17.7 | Y | N | 0.3 | - | - | + | - | - | - | + | - |
| | | 3 | | | M | 12-17 | 19.1 | | | - | 95.9 | - | + | - | - | - | + | - |
| | | 6 | | | M | 12-17 | 19.5 | | | - | 85.7 | - | + | - | - | - | + | - |
| | | 20 | | | M | 12-17 | 19.6 | | | - | 98 | - | + | - | - | - | + | + |
| 336 | pwCF | B | F508del | 1138INSG | M | 31-50 | 28.2 | Y | Y | - | 55 | - | - | + | - | - | - | - |
| | | 3 | | | M | 31-50 | 29.3 | | | - | 63.9 | - | - | + | - | - | - | - |
| | | 6¥ | | | M | 31-50 | 29.0 | | | - | 64 | - | - | + | - | - | - | - |
| 398 | pwCF | B | F508del | 1507del | F | 12-17 | 17.8 | Y | N | - | 101 | - | - | + | - | - | - | - |

| | | | | | | | | | | | | | | | | | | |
|---|---|---|---|---|---|---|---|---|---|---|---|---|---|---|---|---|---|---|
| | | 3 | | | F | 12-17 | 18.6 | | | - | 115 | - | - | + | - | - | - | - |
| | | 6 | | | F | 12-17 | 18.4 | | | - | 100 | - | - | + | - | - | - | - |
| | | 19 | | | F | 12-17 | 20.6 | | | - | 102 | - | - | + | - | - | - | - |
| 437 | pwCF | B* | F508del | F508del | M | 12-17 | 20.5 | Y | N | - | - | - | - | + | - | - | - | - |
| | | 3 | | | M | 12-17 | 21.3 | | | - | 98 | - | - | + | - | - | - | - |
| | | 6 | | | M | 12-17 | 21.2 | | | - | 99.4 | - | - | + | - | - | - | - |
| | | 19 | | | M | 18-23 | 21.3 | | | - | 104 | - | - | + | - | - | - | - |
| 503 | pwCF | B | F508del | F508del | F | 24-30 | 21.7 | Y | N | 32.0 | 57 | - | - | - | - | - | - | - |
| | | 3 | | | F | 24-30 | 23.1 | | | - | 83.3 | - | - | - | - | - | - | - |
| | | 6* | | | F | 24-30 | 23.6 | | | - | 84.3 | - | - | - | - | - | - | - |
| 559 | pwCF | B | F508del | F508del | M | 12-17 | 20.4 | Y | N | - | - | - | - | + | - | + | - | - |
| | | 6 | | | M | 18-23 | 21.4 | | | <5 | 106 | - | - | + | - | - | - | - |
| 696 | pwCF | B | F508del | 711+1G->T, c579+1G>T | M | 12-17 | 16.9 | Y | N | - | 95 | - | - | + | - | - | - | - |
| | | 3 | | | M | 12-17 | 18.1 | | | - | 93 | - | - | + | - | - | - | - |
| | | 6 | | | M | 12-17 | 17.7 | | | - | 93.3 | - | - | + | - | - | - | - |
| 741 | pwCF | B | F508del | 1507del | M | 12-17 | 14.7 | Y | N | <5 | 58.3 | + | - | + | - | - | - | - |
| | | 3 | | | M | 12-17 | 15.7 | | | <5 | 77.9 | + | - | + | - | - | - | - |
| | | 6 | | | M | 12-17 | 15.5 | | | <5 | 72 | + | - | + | - | - | - | - |
| 752 | pwCF | B | F508del | F508del | M | 18-23 | 20.8 | Y | N | - | 116 | - | - | - | - | - | - | - |
| | | 3 | | | M | 18-23 | 21.1 | | | - | - | - | - | - | - | - | - | - |
| | | 6 | | | M | 18-23 | 21.0 | | | - | 115.5 | - | - | - | - | - | - | - |
| | | 23 | | | M | 18-23 | 22.9 | | | - | 118 | - | - | - | - | - | - | - |
| 756 | pwCF | 3 | F508del | 1154insTC, p.Phe342HisfsX28 | F | 12-17 | 22.6 | Y | N | - | 66 | - | - | + | - | - | - | - |
| | | 6 | | | F | 12-17 | 22.2 | | | - | 80.4 | - | - | + | - | - | - | - |
| 802 | pwCF | B | F508del | F508del | M | 18-23 | 20.6 | Y | N | 7.4 | 99 | - | - | - | - | + | - | - |
| | | 3 | | | M | 18-23 | 25.3 | | | 8.0 | - | - | - | - | - | - | - | - |
| | | 6 | | | M | 18-23 | 26.2 | | | 8.0 | 111.7 | - | - | - | - | - | - | - |
| 820 | pwCF | B | F508del | F508del | M | 31-50 | 25.2 | Y | Y | - | 60 | + | - | + | - | + | - | - |

| | | | | | | | | | | | | | | | | | | |
|---|---|---|---|---|---|---|---|---|---|---|---|---|---|---|---|---|---|---|
| | | 3 | | | M | 31-50 | 27.3 | | | 6.0 | 76.8 | + | - | + | - | + | - | - |
| | | 6 | | | M | 31-50 | 28.1 | | | 6.0 | 73.6 | + | - | + | - | + | - | - |
| | | 17 | | | M | 31-50 | 28.2 | | | - | 79.3 | + | - | + | - | + | - | - |
| 868 | pwCF | B | F508del | F508del | M | 24-30 | 18.5 | Y | N | 33.2 | - | + | - | - | + | - | - | - |
| | | 3 | | | M | 24-30 | 20.6 | | | - | - | + | - | - | + | - | - | - |
| | | 6 | | | M | 24-30 | 19.6 | | | - | 67.3 | + | - | - | + | - | - | - |
| 871 | pwCF | B | F508del | gLn1291his | F | 31-50 | 25.9 | Y | N | 29.0 | 75 | - | - | - | - | - | - | - |
| | | 3 | | | F | 31-50 | 27.2 | | | - | - | - | - | - | - | - | - | - |
| 884 | pwCF | B | F508del | F508del | M | 31-50 | 26.3 | Y | Y | - | 58.7 | - | - | + | + | - | - | - |
| | | 3 | | | M | 31-50 | 27.4 | | | 3.4 | 67.4 | - | - | + | + | - | - | - |
| | | 6 | | | M | 31-50 | 28.2 | | | 3.4 | 71.4 | - | - | + | + | - | - | - |
| | | 19 | | | M | 31-50 | 27.8 | | | - | 68 | - | - | + | + | - | - | - |
| 152 | HC | - | Unknown | Unknown | F | 18-23 | 21.3 | N | - | 4.2 | - | - | - | - | - | - | - | - |
| 159 | HC | - | Unknown | Unknown | M | 12-17 | 23.5 | N | - | 2.7 | - | - | - | - | - | - | - | - |
| 205 | HC | - | Unknown | Unknown | F | 18-23 | 31.9 | N | - | 3.8 | - | - | - | - | - | - | - | - |
| 431 | HC | - | Unknown | Unknown | M | 12-17 | 18.0 | N | - | 2.4 | - | - | - | - | - | - | - | - |
| 501 | HC | - | Unknown | Unknown | F | 24-30 | 28.7 | N | - | 7.2 | - | - | - | - | - | - | - | - |
| 548 | HC | - | Unknown | Unknown | M | 24-30 | 24.5 | N | - | 3.6 | - | - | - | - | - | - | - | - |
| 673 | HC | - | Unknown | Unknown | M | 18-23 | 20.3 | N | - | 0.9 | - | - | - | - | - | - | - | - |
| 749 | HC | - | Unknown | Unknown | F | 31-50 | 19.6 | N | - | 3.0 | - | - | - | - | - | - | - | - |
| 964 | HC | - | Unknown | Unknown | F | 12-17 | 19.2 | N | - | 12.7 | - | - | - | - | - | - | - | - |
| 986 | HC | - | Unknown | Unknown | M | 24-30 | 22.6 | N | - | 5.0 | - | - | - | - | - | - | - | - |

Treatment length points marked indicate samples for which no *metagenomic or ¥metabolomic analysis was available, due incomplete stool sampling or following removal of reads/data from downstream processing. CF participants were either F508del homozygous, or F508del heterozygous. All patients were pancreatic insufficient but contained no CF-related diabetes. For antibiotic usage, '+' indicates routine use of antibiotic class during the period for which the respective sample was obtained. Abbreviations: pwCF - People with CF, HC - Healthy controls, B - Baseline, FEV1 – Percent predicted forced expiratory volume in 1 second, BMI – Body mass index, PI – Pancreatic insufficiency, CFRD – Cystic fibrosis-related diabetes, FCP - Faecal calprotectin, Antibiotics; Ami – Aminoglycosides, Ant – Antimycobacterials, Mac – Macrolides, Mon – Monobactams, Pol – Polymixins, Sul – Sulfonamides, Tet – Tetracyclines.

**Table S2**. Magnetic resonance imaging (MRI) data between pwCF and healthy control participants.

| Participant ID | Group | Treatment length (months) | OCTT (mins) | SBWC ($mL/m^2$) |
|---|---|---|---|---|
| 023 | pwCF | B | >360 | 54.10 |
| | | 3 | >360 | 71.45 |
| | | 6 | >360 | 83.34 |
| | | 20 | 240 | 43.08 |
| 046 | pwCF | B | 120 | 83.35 |
| | | 3* | >360 | 49.31 |
| | | 6 | >360 | 56.03 |
| 194 | pwCF | B | >360 | 79.48 |
| | | 3 | >360 | 119.20 |
| | | 6 | >360 | 72.16 |
| 247 | pwCF | B | 180 | 273.09 |
| | | 3 | 300 | 62.29 |
| 253 | pwCF | B | >360 | 82.92 |
| | | 3 | >360 | 74.79 |
| | | 6 | >360 | 82.09 |
| 257 | pwCF | B | 240 | 40.56 |
| | | 3 | >360 | 53.51 |
| | | 6 | 120 | 20.44 |
| | | 20 | 120 | 58.45 |
| 336 | pwCF | B | >360 | 45.65 |
| | | 3 | >360 | 51.95 |
| | | 6 | >360 | 57.06 |
| 398 | pwCF | B | >360 | 136.36 |
| | | 3 | 240 | 180.52 |
| | | 6 | 240 | 102.51 |
| | | 19 | 360 | 169.15 |
| 437 | pwCF | B* | >360 | 99.35 |
| | | 3 | >360 | 93.81 |
| | | 6 | 360 | 83.35 |
| | | 19 | 180 | 86.85 |
| 503 | pwCF | B | >360 | 31.25 |
| | | 3 | >360 | 49.21 |
| | | 6* | 240 | 59.00 |
| 559 | pwCF | B | >360 | 74.30 |
| | | 6 | >360 | 95.80 |
| 696 | pwCF | B | 360 | 43.59 |
| | | 3 | 360 | 54.28 |
| | | 6 | 300 | 37.07 |
| 741 | pwCF | B | >360 | 38.34 |

| | | | | |
|---|---|---|---|---|
| | | 3 | 360 | 45.91 |
| | | 6 | >360 | 51.92 |
| 752 | pwCF | B | >360 | 42.38 |
| | | 3 | 300 | 105.59 |
| | | 6 | >360 | 96.71 |
| | | 23 | 180 | 138.62 |
| 756 | pwCF | 3 | >360 | 105.59 |
| | | 6 | >360 | 96.71 |
| 802 | pwCF | B | >360 | 133.05 |
| | | 3 | >360 | 38.02 |
| | | 6 | >360 | 25.13 |
| 820 | pwCF | B | 300 | 41.53 |
| | | 3 | 300 | 59.30 |
| | | 6 | 300 | 49.08 |
| | | 17 | 300 | 54.55 |
| 868 | pwCF | B | >360 | 82.84 |
| | | 3 | 300 | 118.88 |
| | | 6 | 240 | 113.28 |
| 871 | pwCF | B | - | - |
| | | 3 | - | - |
| 884 | pwCF | B | 300 | 97.00 |
| | | 3 | 300 | 47.33 |
| | | 6 | >360 | 58.21 |
| | | 19 | 240 | 59.05 |
| 152 | HC | - | 180 | 61.51 |
| 159 | HC | - | 150 | 18.40 |
| 205 | HC | - | 360 | 40.35 |
| 431 | HC | - | 150 | 72.69 |
| 501 | HC | - | 360 | 47.56 |
| 548 | HC | - | 300 | 57.04 |
| 673 | HC | - | 360 | 24.16 |
| 749 | HC | - | 240 | 10.79 |
| 964 | HC | - | 180 | 26.19 |
| 986 | HC | - | 180 | 89.90 |

B; Baseline, 3; 3 Months ETI therapy, 6; 6 Months ETI therapy, 17+; Extended (17+ months) ETI therapy, OCTT; Oro-caecal transit time, SBWC; Small bowel water content (corrected for body surface area).

**Table S3**. Analytical parameters for SCFA analysis with GC-MS

| SCFA | $t_R$ (min) | Target ion (m/z) | Confirmation ion (m/z) |
|---|---|---|---|
| **Acetic acid** | 2.52 | 117 | 75 |
| **Propionic acid** | 3.81 | 131 | 75 |
| **Iso-butyric acid** | 4.39 | 145 | 117 |
| **Butyric acid** | 5.22 | 145 | 117 |
| **Iso-valeric acid** | 6.07 | 159 | 117 |
| **Valeric acid** | 6.85 | 159 | 117 |
| **4-methylvaleric acid** | 7.86 | 173 | 117 |
| **Hexanoic acid** | 8.41 | 173 | 117 |
| **Heptanoic acid** | 9.9 | 187 | 117 |
| **$^{13}$C-acetic acid** | 2.52 | 119 | 77 |
| **$^{13}$C-propionic acid** | 3.81 | 134 | 77 |
| **$^{13}$C-butyric acid** | 5.22 | 149 | 119 |

$t_R$ - Retention time

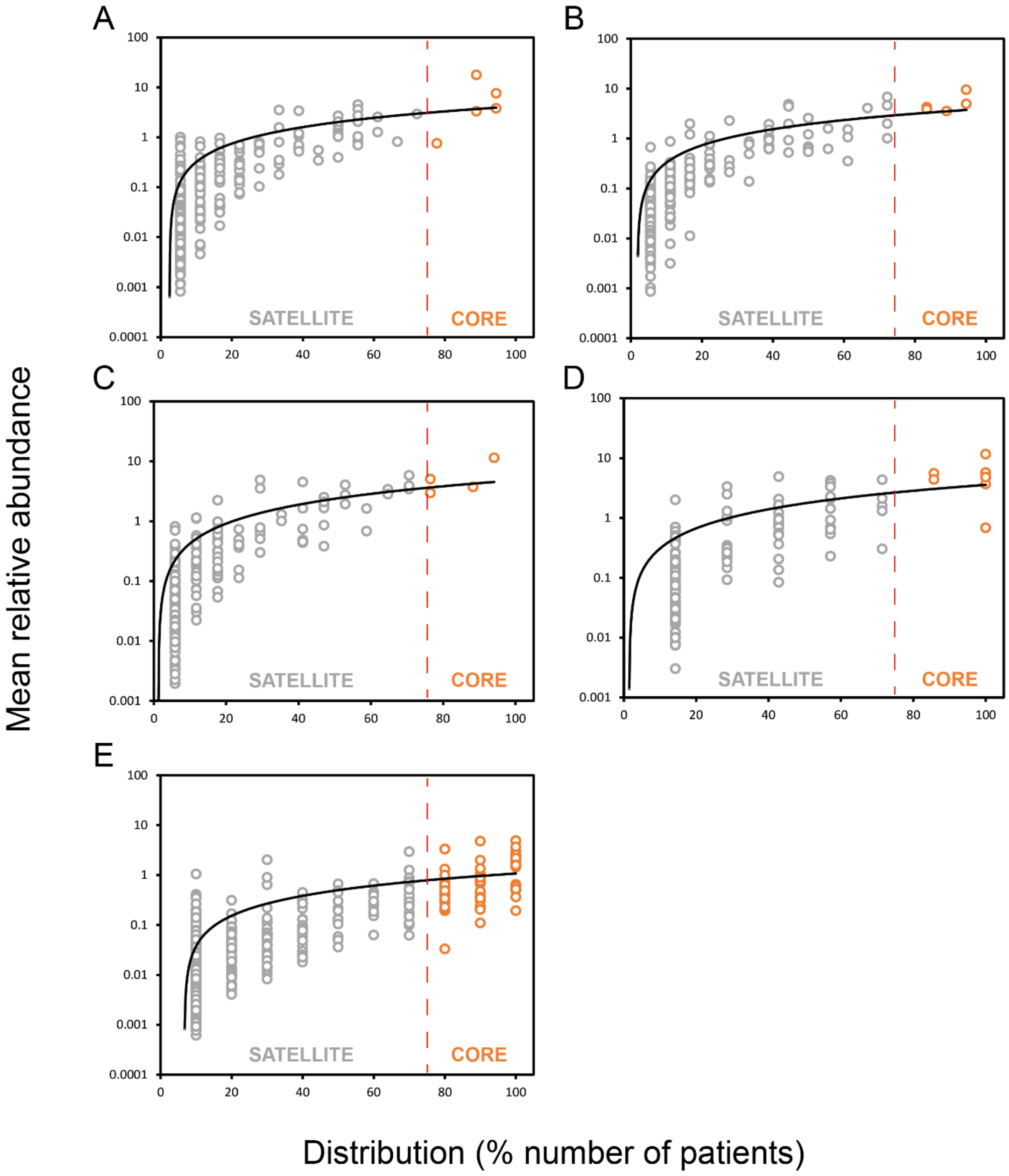

**Figure S1**. Distribution and abundance of bacterial taxa across lengthening ETI treatment (0, 3, 6, 17+ months) stages (A, B, C, D respectively) and healthy control participants (E). Given is the percentage number of patients harbouring each bacterial taxon, plotted against the mean relative abundance across those samples. Core taxa are depicted by the orange circles and fall in the upper quartile of distribution, separated by the red vertical line at 75% distribution. Satellite taxa (grey) are all samples below this distribution. Distribution-abundance relationship regression statistics: (a) $r^2$ = 0.45, $F_{1,320}$ = 249.4, P < 0.0001. (b) $r^2$ = 0.63, $F_{1,253}$ = 434.1, P < 0.0001. (c) $r^2$ = 0.65, $F_{1,196}$ = 358.5, P < 0.0001. (d) $r^2$ = 0.53, $F_{1,146}$ = 167.2, P < 0.0001. (e) $r^2$ = 0.40, $F_{1,482}$ = 321.4, P < 0.0001.

**Table S4**. Core taxa throughout ETI therapy and control participants.

| | Mean relative abundance (%) | | | | |
|---|---|---|---|---|---|
| | ETI duration (months) | | | | |
| *Genus* | 0 | 3 | 6 | 17+ | HC |
| *Blautia 1* | 17.69 | 9.54 | 11.45 | 11.66 | 4.91 |
| *Blautia luti* | 4.49 | 6.79 | 5.83 | 5.55 | 3.01 |
| *Fusicatenibacter saccharivorans* | 7.61 | 4.24 | 2.96 | 5.73 | 3.69 |
| *Dorea longicatena* | 3.61 | 4.61 | 5.05 | 4.39 | 1.79 |
| *Eubacterium hallii* | 3.79 | 3.56 | 3.73 | 4.81 | 2.39 |
| *Bacteroides dorei* | 2.67 | 2.56 | 4.05 | 3.79 | 3.35 |
| *Anaerostipes hadrus* | 2.73 | 4.06 | 2.83 | 4.35 | 1.99 |
| *Eubacterium rectale* | 1.39 | 2.55 | 3.43 | 3.68 | 4.78 |
| *Collinsella aerofaciens* | 3.37 | 4.85 | 3.52 | 3.33 | 0.19 |
| *Gemmiger formicilis* | 1.05 | 4.40 | 4.51 | 4.24 | 1.05 |
| *Clostridium disporicum* | 2.92 | 4.97 | 3.90 | 1.60 | 0.70 |
| *Bifidobacterium adolescentis* | 0.82 | 2.29 | 4.88 | 4.90 | 0.81 |
| *Streptococcus 6* | 3.32 | 3.87 | 3.40 | 1.43 | 0.19 |
| *Blautia faecis* | 2.53 | 1.98 | 1.94 | 2.12 | 2.30 |
| *Bacteroides vulgatus* | 1.57 | 0.52 | 2.23 | 1.66 | 3.32 |
| *Escherichia coli* | 2.05 | 0.70 | 1.66 | 2.27 | 1.99 |
| *Romboutsia timonensis* | 1.16 | 1.96 | 2.75 | 0.76 | 1.32 |
| *Ruminococcus faecis* | 1.20 | 0.96 | 1.64 | 2.27 | 1.61 |
| *Blautia obeum* | 1.75 | 1.07 | 0.73 | 0.98 | 2.51 |
| *Faecalibacterium duncaniae* | 0.49 | 0.86 | 0.52 | 2.09 | 2.15 |
| *Intestinibacter bartlettii* | 0.34 | 1.07 | 1.01 | 1.22 | 0.63 |
| *Bacteroides uniformis* | 0.66 | 0.49 | 0.41 | 0.14 | 2.55 |
| *Eubacterium desmolans* | 0.76 | 1.01 | 0.68 | 0.69 | 0.67 |
| *Faecalibacillus 70* | 0.18 | 0.78 | 0.70 | 0.54 | 1.45 |
| *Dorea phocaeensis* | 0.27 | 0.68 | 0.86 | 1.31 | 0.31 |
| *Coprococcus comes* | 0.52 | 0.52 | 0.44 | 0.92 | 0.57 |
| *Faecalibacterium longum* | 0.03 | 0.14 | 0.15 | 0.42 | 2.19 |
| *Parabacteroides distasonis* | 1.04 | 0.21 | 0.15 | 0.11 | 0.90 |
| *Faecalibacterium prausnitzii* | 0.14 | 0.00 | 0.03 | 0.00 | 2.04 |
| *Roseburia hominis* | 0.60 | 0.24 | 0.22 | 0.00 | 1.00 |
| *Blautia intestinalis* | 0.29 | 0.27 | 0.58 | 0.00 | 0.50 |
| *Gemmiger 86* | 0.03 | 0.17 | 0.21 | 0.47 | 0.75 |
| *Barnesiella intestinihominis* | 0.01 | 0.03 | 0.01 | 0.00 | 1.58 |
| *Hominisplanchenecus faecis* | 0.49 | 0.31 | 0.03 | 0.14 | 0.50 |
| *Alistipes putredinis* | 0.00 | 0.00 | 0.00 | 0.00 | 1.35 |
| *Eubacterium coprostanoligenes* | 0.04 | 0.39 | 0.04 | 0.27 | 0.43 |
| *Blautia torques* | 0.21 | 0.04 | 0.07 | 0.30 | 0.39 |
| *Clostridium 146* | 0.00 | 0.27 | 0.03 | 0.18 | 0.43 |
| *Eubacterium ramulus* | 0.14 | 0.17 | 0.07 | 0.32 | 0.21 |
| *Ruminococcus champanellensis* | 0.00 | 0.02 | 0.00 | 0.21 | 0.65 |

| | | | | | |
|---|---|---|---|---|---|
| *Lachnoclostridium edouardi* | 0.09 | 0.15 | 0.04 | 0.00 | 0.53 |
| *Alistipes onderdonkii* | 0.04 | 0.00 | 0.01 | 0.00 | 0.64 |
| *Eubacterium ventriosum* | 0.03 | 0.05 | 0.04 | 0.26 | 0.29 |
| *Bacteroides caccae* | 0.03 | 0.03 | 0.15 | 0.06 | 0.36 |
| *Odoribacter splanchnicus* | 0.01 | 0.00 | 0.00 | 0.00 | 0.60 |
| *Coprococcus catus* | 0.08 | 0.07 | 0.02 | 0.14 | 0.22 |
| *Waltera intestinalis* | 0.02 | 0.00 | 0.00 | 0.00 | 0.49 |
| *Oscillibacter 413* | 0.00 | 0.00 | 0.00 | 0.00 | 0.48 |
| *Evtepia gabavorous* | 0.00 | 0.00 | 0.00 | 0.00 | 0.47 |
| *Vescimonas fastidiosa* | 0.00 | 0.03 | 0.00 | 0.00 | 0.41 |
| *Lentihominibacter faecis* | 0.06 | 0.08 | 0.07 | 0.08 | 0.11 |
| *Roseburia inulinivorans* | 0.05 | 0.00 | 0.00 | 0.00 | 0.34 |
| *Marseillibacter massiliensis* | 0.00 | 0.03 | 0.00 | 0.00 | 0.34 |
| *Alistipes obesi* | 0.00 | 0.00 | 0.00 | 0.00 | 0.36 |
| *Alistipes marseilloanorexicus* | 0.02 | 0.00 | 0.00 | 0.00 | 0.32 |
| *Eubacterium eligens* | 0.00 | 0.05 | 0.00 | 0.00 | 0.29 |
| *Alistipes shahii* | 0.00 | 0.00 | 0.00 | 0.00 | 0.24 |
| *Vescimonas 720* | 0.00 | 0.00 | 0.00 | 0.00 | 0.23 |
| *Oscillibacter massiliensis* | 0.00 | 0.01 | 0.01 | 0.00 | 0.20 |
| *Pseudomonas 1352* | 0.01 | 0.00 | 0.00 | 0.00 | 0.03 |

175

**Table S5**. Kruskal-Wallis tests of bacterial alpha diversity across treatment time-points (months) utilising Fisher's alpha index.

| | **Microbiota** | | **Core** | | **Satellite** | |
|---|---|---|---|---|---|---|
| 0-3 | K (Observed value) | 1.851 | K (Observed value) | 0.169 | K (Observed value) | 1.894 |
| | K (Critical value) | 3.841 | K (Critical value) | 3.841 | K (Critical value) | 3.841 |
| | DF | 1 | DF | 1 | DF | 1 |
| | p-value (one-tailed) | 0.174 | p-value (one-tailed) | 0.681 | p-value (one-tailed) | 0.169 |
| 0-6 | K (Observed value) | 12.240 | K (Observed value) | 9.021 | K (Observed value) | 11.333 |
| | K (Critical value) | 3.841 | K (Critical value) | 3.841 | K (Critical value) | 3.841 |
| | DF | 1 | DF | 1 | DF | 1 |
| | p-value (one-tailed) | 0.0005 | p-value (one-tailed) | 0.003 | p-value (one-tailed) | 0.001 |
| 0-17+ | K (Observed value) | 0.938 | K (Observed value) | 14.538 | K (Observed value) | 1.059 |
| | K (Critical value) | 3.841 | K (Critical value) | 3.841 | K (Critical value) | 3.841 |
| | DF | 1 | DF | 1 | DF | 1 |
| | p-value (one-tailed) | 0.333 | p-value (one-tailed) | 0.00014 | p-value (one-tailed) | 0.304 |
| 3-6 | K (Observed value) | 4.462 | K (Observed value) | 9.938 | K (Observed value) | 2.946 |
| | K (Critical value) | 3.841 | K (Critical value) | 3.841 | K (Critical value) | 3.841 |
| | DF | 1 | DF | 1 | DF | 1 |
| | p-value (one-tailed) | 0.035 | p-value (one-tailed) | 0.002 | p-value (one-tailed) | 0.086 |
| 6-17+ | K (Observed value) | 2.623 | K (Observed value) | 14.280 | K (Observed value) | 1.614 |
| | K (Critical value) | 3.841 | K (Critical value) | 3.841 | K (Critical value) | 3.841 |
| | DF | 1 | DF | 1 | DF | 1 |
| | p-value (one-tailed) | 0.105 | p-value (one-tailed) | 0.00016 | p-value (one-tailed) | 0.204 |

**Table S6**. Kruskal-Wallis tests of bacterial alpha diversity between treatment time-points (months) and healthy controls utilising Fisher's alpha index.

| | **Microbiota** | | **Core** | | **Satellite** | |
|---|---|---|---|---|---|---|
| 0-HC | K (Observed value) | 18.621 | K (Observed value) | 18.621 | K (Observed value) | 18.626 |
| | K (Critical value) | 3.841 | K (Critical value) | 3.841 | K (Critical value) | 3.841 |
| | DF | 1 | DF | 1 | DF | 1 |
| | p-value (one-tailed) | <0.0001 | p-value (one-tailed) | <0.0001 | p-value (one-tailed) | <0.0001 |
| 3-HC | K (Observed value) | 18.621 | K (Observed value) | 18.626 | K (Observed value) | 18.626 |
| | K (Critical value) | 3.841 | K (Critical value) | 3.841 | K (Critical value) | 3.841 |
| | DF | 1 | DF | 1 | DF | 1 |
| | p-value (one-tailed) | <0.0001 | p-value (one-tailed) | <0.0001 | p-value (one-tailed) | <0.0001 |
| 6-HC | K (Observed value) | 18.214 | K (Observed value) | 18.214 | K (Observed value) | 18.220 |
| | K (Critical value) | 3.841 | K (Critical value) | 3.841 | K (Critical value) | 3.841 |
| | DF | 1 | DF | 1 | DF | 1 |
| | p-value (one-tailed) | <0.0001 | p-value (one-tailed) | <0.0001 | p-value (one-tailed) | <0.0001 |
| 17+-HC | K (Observed value) | 11.667 | K (Observed value) | 11.667 | K (Observed value) | 11.681 |
| | K (Critical value) | 3.841 | K (Critical value) | 3.841 | K (Critical value) | 3.841 |
| | DF | 1 | DF | 1 | DF | 1 |
| | p-value (one-tailed) | 0.001 | p-value (one-tailed) | 0.001 | p-value (one-tailed) | 0.001 |

**Table S7**. Bacterial ANOSIM summary statistics across ETI treatment lengths, utilising the Bray-Curtis index.

| | Microbiota | | Core | | Satellite | |
|---|---|---|---|---|---|---|
| 0-3 | R | -0.02372 | R | 0.02923 | R | 0.0382 |
| | p (same) | 0.7567 | p (same) | 0.1471 | p (same) | 0.1355 |
| | Bonferroni-corrected p value | 0.7598 | Bonferroni-corrected p value | 0.1481 | Bonferroni-corrected p value | 0.1421 |
| | Permutations | 9999 | Permutations | 9999 | Permutations | 9999 |
| 0-6 | R | -0.03417 | R | 0.1835 | R | 0.03914 |
| | p (same) | 0.8365 | p (same) | 0.0001 | p (same) | 0.1395 |
| | Bonferroni-corrected p value | 0.8495 | Bonferroni-corrected p value | 0.0002 | Bonferroni-corrected p value | 0.1477 |
| | Permutations | 9999 | Permutations | 9999 | Permutations | 9999 |
| 0-17+ | R | -0.1927 | R | 0.1942 | R | 0.09095 |
| | p (same) | 0.9837 | p (same) | 0.0547 | p (same) | 0.1684 |
| | Bonferroni-corrected p value | 0.985 | Bonferroni-corrected p value | 0.0614 | Bonferroni-corrected p value | 0.1763 |
| | Permutations | 9999 | Permutations | 9999 | Permutations | 9999 |
| 3-6 | R | -0.0664 | R | 0.2078 | R | 0.07737 |
| | p (same) | 0.9938 | p (same) | 0.0001 | p (same) | 0.034 |
| | Bonferroni-corrected p value | 0.9947 | Bonferroni-corrected p value | 0.0001 | Bonferroni-corrected p value | 0.0347 |
| | Permutations | 9999 | Permutations | 9999 | Permutations | 9999 |
| 6-17+ | R | -0.1876 | R | 0.1817 | R | 0.04437 |
| | p (same) | 0.9798 | p (same) | 0.0591 | p (same) | 0.288 |
| | Bonferroni-corrected p value | 0.9806 | Bonferroni-corrected p value | 0.0639 | Bonferroni-corrected p value | 0.2944 |
| | Permutations | 9999 | Permutations | 9999 | Permutations | 9999 |

**Table S8**. Bacterial ANOSIM summary statistics between ETI treatment lengths and healthy control participants, utilising the Bray-Curtis index.

| | Microbiota | | Core | | Satellite | |
|---|---|---|---|---|---|---|
| 0-HC | R | 0.2775 | R | 0.7931 | R | 0.8795 |
| | p (same) | 0.0018 | p (same) | 0.0001 | p (same) | 0.0001 |
| | Bonferroni-corrected p value | 0.0023 | Bonferroni-corrected p value | 0.0001 | Bonferroni-corrected p value | 0.0001 |
| | Permutations | 9999 | Permutations | 9999 | Permutations | 9999 |
| 3-HC | R | 0.3104 | R | 0.7432 | R | 0.8999 |
| | p (same) | 0.0015 | p (same) | 0.0001 | p (same) | 0.0001 |
| | Bonferroni-corrected p value | 0.0021 | Bonferroni-corrected p value | 0.0001 | Bonferroni-corrected p value | 0.0001 |
| | Permutations | 9999 | Permutations | 9999 | Permutations | 9999 |
| 6-HC | R | 0.3751 | R | 0.9177 | R | 0.9299 |
| | p (same) | 0.0002 | p (same) | 0.0001 | p (same) | 0.0001 |
| | Bonferroni-corrected p value | 0.0002 | Bonferroni-corrected p value | 0.0001 | Bonferroni-corrected p value | 0.0001 |
| | Permutations | 9999 | Permutations | 9999 | Permutations | 9999 |
| 17+-HC | R | 0.6697 | R | 0.9957 | R | 0.8723 |
| | p (same) | 0.0002 | p (same) | 0.0001 | p (same) | 0.0001 |
| | Bonferroni-corrected p value | 0.0002 | Bonferroni-corrected p value | 0.0002 | Bonferroni-corrected p value | 0.0002 |
| | Permutations | 9999 | Permutations | 9999 | Permutations | 9999 |

179

**Table S9**. Similarity of percentage (SIMPER) analysis of microbiota dissimilarity (Bray-Curtis) between baseline and 17+ months (extended) ETI treatment in samples obtained from pwCF.

| Taxa | % Relative abundance | | Av. Dissimilarity | % Contribution | Cumulative (%) |
|---|---|---|---|---|---|
| | Baseline | 17+ ETI | | | |
| *Blautia 1* | 17.70 | 11.70 | 5.59 | 7.62 | 7.621 |
| *Fusicatenibacter saccharivorans* | 7.61 | 5.73 | 3.56 | 4.85 | 12.47 |
| *Blautia luti* | 4.49 | 5.55 | 2.48 | 3.38 | 15.85 |
| *Bifidobacterium adolescentis* | 0.82 | 4.90 | 2.31 | 3.14 | 18.99 |
| *Collinsella aerofaciens* | 3.37 | 3.33 | 2.27 | 3.10 | 22.09 |
| *Anaerostipes hadrus* | 2.73 | 4.35 | 2.24 | 3.05 | 25.14 |
| *Bacteroides dorei* | 2.67 | 3.79 | 2.21 | 3.01 | 28.15 |
| *Dorea longicatena* | 3.61 | 4.39 | 2.01 | 2.74 | 30.89 |
| *Gemmiger formicilis* | 1.05 | 4.24 | 1.82 | 2.48 | 33.36 |
| *Bifidobacterium 21* | 3.50 | 0.22 | 1.67 | 2.27 | 35.64 |
| *Enterococcus 26* | 2.55 | 2.07 | 1.54 | 2.10 | 37.74 |
| *Eubacterium rectale* | 1.39 | 3.68 | 1.53 | 2.09 | 39.82 |
| *Ruminococcus bromii* | 0.16 | 3.34 | 1.48 | 2.02 | 41.84 |
| *Streptococcus 6* | 3.32 | 1.43 | 1.43 | 1.95 | 43.79 |
| *Eubacterium hallii* | 3.79 | 4.81 | 1.37 | 1.87 | 45.66 |
| *Clostridium disporicum* | 2.92 | 1.60 | 1.37 | 1.87 | 47.53 |
| *Ruminococcus gnavus* | 1.26 | 2.48 | 1.33 | 1.81 | 49.34 |
| *Escherichia coli* | 2.05 | 2.27 | 1.29 | 1.75 | 51.09 |

Taxa identified as core are highlighted in orange, with satellite taxa highlighted in grey. The mean relative abundance (%) across both groups is given, alongside the percentage contribution which is the mean dissimilarity of taxa divided by the mean dissimilarity (73.40%) across samples. Cumulative percent does not equal 100% as the list is not exhaustive, rather the taxa that make up 50% of dissimilarity between groups. Given the length of the 16S gene regions sequenced, taxon identification should be considered putative.

**Table S10**. Redundancy analysis to explain percent variation across whole microbiota, core, and satellite taxa of the significant clinical variables across pwCF receiving ETI therapy.

| | Microbiota | | | Core taxa | | | Satellite taxa | | |
|---|---|---|---|---|---|---|---|---|---|
| | Var. Exp (%) | pseudo-*F* | *P* (adj) | Var. Exp (%) | pseudo-*F* | *P* (adj) | Var. Exp (%) | pseudo-*F* | *P* (adj) |
| Age | | | | | | | 6.5 | 3.4 | 0.002 |
| Antibiotics | 4.9 | 2.8 | 0.010 | 5.6 | 3.3 | 0.050 | 5.4 | 2.9 | 0.002 |
| BMI | 5.2 | 2.9 | 0.016 | | | | | | |
| Disease sev. | 7.4 | 3.9 | 0.002 | 9.6 | 5.2 | 0.006 | 3.4 | 1.9 | 0.012 |
| SBWC | 3.4 | 2.0 | 0.046 | 4.9 | 3.0 | 0.038 | | | |
| Sex | 3.7 | 2.2 | 0.038 | 5.5 | 3.1 | 0.082 | 2.6 | 1.5 | 0.050 |
| Treatment length | | | | | | | 2.9 | 1.7 | 0.048 |
| Total | **24.6** | | | **25.6** | | | **20.8** | | |

Var. Exp (%) represents the percentage of the microbiota for which can be explained by a given clinical variable from the redundancy analysis model. *P* (adj) is following false discovery rate correction of significance values. Antibiotics refers to if a given participant was receiving any class of routine antibiotic during the sampling period. Disease sev – Disease severity utilising FEV1% as a proxy, SBWC – Small bowel water content corrected for body surface area.

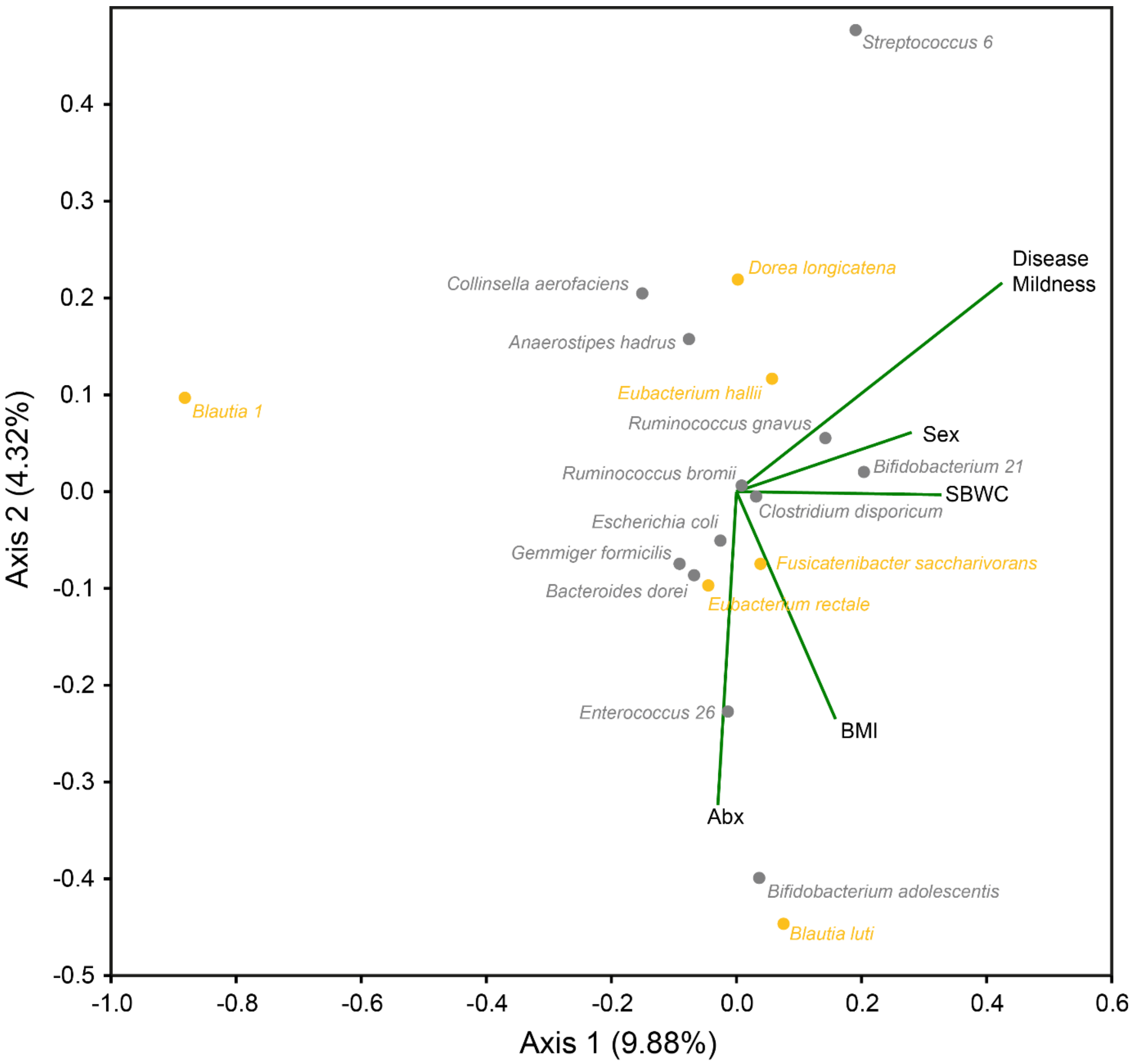

**Figure S2**. Redundancy analysis species biplots for the whole microbiota within pwCF. The 24 taxa contributing most to the dissimilarity (cumulatively > 50%) within pwCF samples at baseline, and following 17+ months (extended) ETI therapy from the SIMPER analysis (Table S9) are shown independently of the total number of ASVs identified (353). Orange points represent taxa that were identified as core for the pwCF group following 17+ (extended) ETI therapy, grey points are satellite, and the white (black stroke) points represent taxa that were absent. Biplot lines depict clinical variables that significantly account for total variation in taxa relative abundance within whole microbiota analysis at the $p \leq 0.05$ level (Table S10). Species plots depict the strength of explanation provided by the given clinical variables, with taxa shown in the same direction of a particular clinical variable considered to have a higher value than those that are not. ‘Abx’ – Antibiotics during sampling period, ‘BMI’ – Body mass index, ‘Disease Mildness’ – Disease mildness based on increased FEV1% across patients, ‘SBWC’ – Small bowel water content corrected for body surface area. The percentage of microbiota variation explained by each axis is given in parentheses.

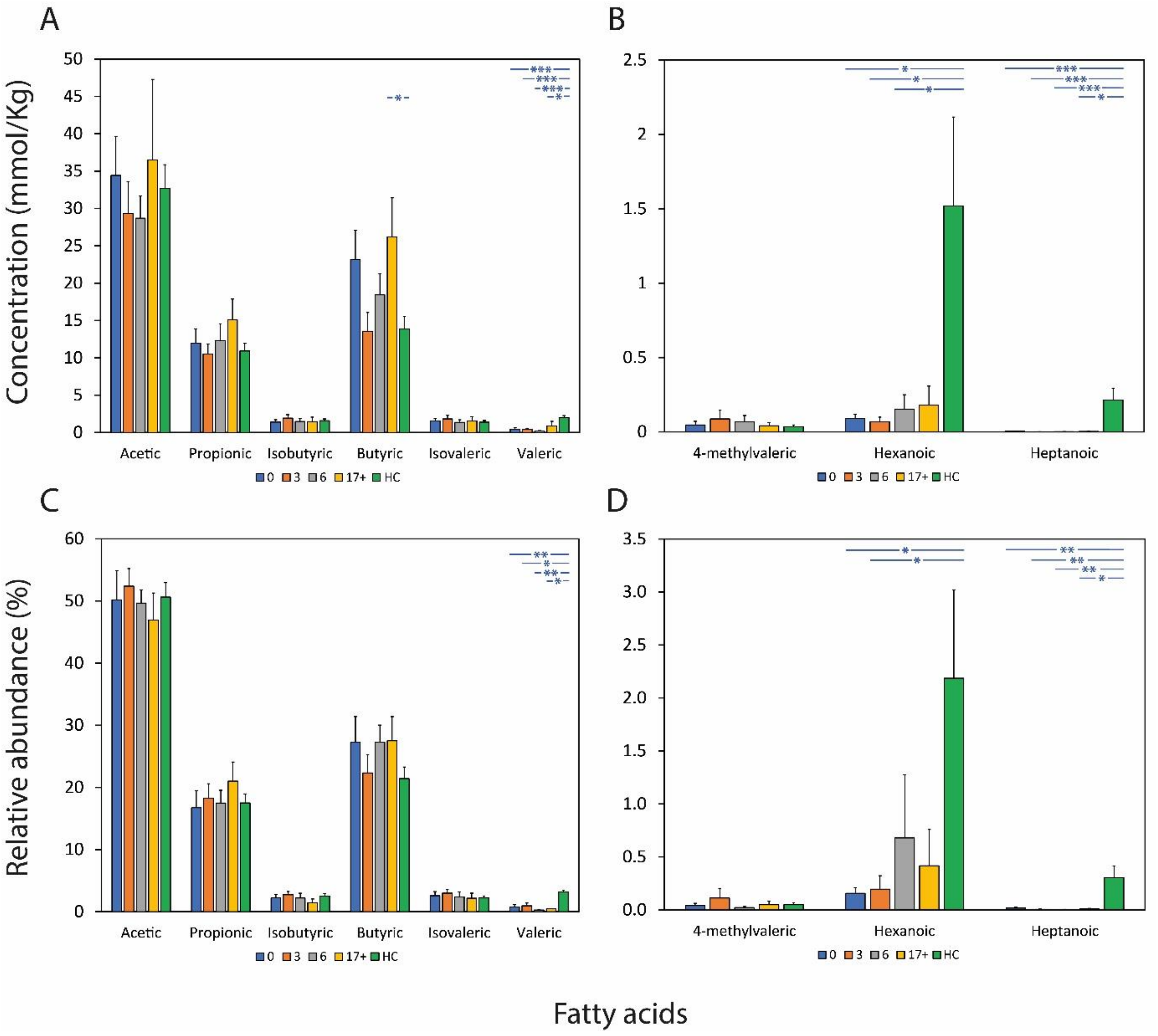

**Figure S3**. Changes in faecal fatty acid concentration (A-B) and relative abundance (C-D) across ETI treatment periods (months) and healthy control samples. Error bars denote standard error of the mean (SEM). Asterisks between bars denote significance differences in absolute quantification (A-B) or relative abundance (C-D) of faecal fatty acids between groups following Kruskal-Wallis testing. ***; *P* < 0.0001, **; *P* < 0.001, *; *P* < 0.05. Summary statistics are found in Tables S11-14.

**Table S11**. Kruskal-Wallis tests of faecal fatty acid quantification between across ETI treatment time-points.

| | | Fatty acid | | | | | | | | |
|---|---|---|---|---|---|---|---|---|---|---|
| | | Acetic | Propionic | Isobutyric | Butyric | Isovaleric | Valeric | 4-methyl | Hexanoic | Heptanoic |
| 0-3 | K (Observed value) | 0.189 | 0.179 | 0.511 | 1.765 | 0.003 | 0.214 | 0.025 | 1.470 | 2.016 |
| | K (Critical value) | 3.841 | 3.841 | 3.841 | 3.841 | 3.841 | 3.841 | 3.841 | 3.841 | 3.841 |
| | DF | 1 | 1 | 1 | 1 | 1 | 1 | 1 | 1 | 1 |
| | p-value (one-tailed) | 0.6635 | 0.6721 | 0.4749 | 0.1841 | 0.9534 | 0.6440 | 0.8755 | 0.2254 | 0.1556 |
| 3-6 | K (Observed value) | 0.107 | 0.236 | 0.810 | 2.793 | 0.351 | 0.086 | 0.095 | 1.872 | 0.137 |
| | K (Critical value) | 3.841 | 3.841 | 3.841 | 3.841 | 3.841 | 3.841 | 3.841 | 3.841 | 3.841 |
| | DF | 1 | 1 | 1 | 1 | 1 | 1 | 1 | 1 | 1 |
| | p-value (one-tailed) | 0.7442 | 0.6268 | 0.3682 | 0.0947 | 0.5535 | 0.7697 | 0.7579 | 0.1712 | 0.7116 |
| 6-17+ | K (Observed value) | 0.044 | 1.059 | 0.000 | 2.289 | 0.366 | 2.476 | 0.007 | 0.004 | 0.040 |
| | K (Critical value) | 3.841 | 3.841 | 3.841 | 3.841 | 3.841 | 3.841 | 3.841 | 3.841 | 3.841 |
| | DF | 1 | 1 | 1 | 1 | 1 | 1 | 1 | 1 | 1 |
| | p-value (one-tailed) | 0.8330 | 0.3035 | 1.0000 | 0.1303 | 0.5450 | 0.1156 | 0.9354 | 0.9517 | 0.8416 |
| 0-6 | K (Observed value) | 0.653 | 0.000 | 0.029 | 0.267 | 0.316 | 0.651 | 0.061 | 0.059 | 1.428 |
| | K (Critical value) | 3.841 | 3.841 | 3.841 | 3.841 | 3.841 | 3.841 | 3.841 | 3.841 | 3.841 |
| | DF | 1 | 1 | 1 | 1 | 1 | 1 | 1 | 1 | 1 |
| | p-value (one-tailed) | 0.4189 | 1.0000 | 0.8651 | 0.6055 | 0.5740 | 0.4198 | 0.8054 | 0.8078 | 0.2321 |
| 0-17+ | K (Observed value) | 0.011 | 1.271 | 0.007 | 0.188 | 0.021 | 1.322 | 0.013 | 0.442 | 0.268 |
| | K (Critical value) | 3.841 | 3.841 | 3.841 | 3.841 | 3.841 | 3.841 | 3.841 | 3.841 | 3.841 |
| | DF | 1 | 1 | 1 | 1 | 1 | 1 | 1 | 1 | 1 |
| | p-value (one-tailed) | 0.9161 | 0.2596 | 0.9343 | 0.6646 | 0.8851 | 0.2502 | 0.9076 | 0.5061 | 0.6049 |

210

**Table S12**. Kruskal-Wallis tests of fatty acid relative abundance between across ETI treatment time-points.

| | | Fatty acid | | | | | | | | |
|---|---|---|---|---|---|---|---|---|---|---|
| | | Acetic | Propionic | Isobutyric | Butyric | Isovaleric | Valeric | 4-methyl | Hexanoic | Heptanoic |
| 0-3 | K (Observed value) | 0.240 | 0.240 | 0.296 | 0.500 | 0.214 | 0.145 | 0.279 | 0.359 | 0.465 |
| | K (Critical value) | 3.841 | 3.841 | 3.841 | 3.841 | 3.841 | 3.841 | 3.841 | 3.841 | 3.841 |
| | DF | 1 | 1 | 1 | 1 | 1 | 1 | 1 | 1 | 1 |
| | p-value (one-tailed) | 0.6245 | 0.6245 | 0.5865 | 0.4795 | 0.6438 | 0.7034 | 0.5976 | 0.5489 | 0.4951 |
| 3-6 | K (Observed value) | 0.068 | 0.303 | 0.706 | 1.854 | 0.568 | 0.806 | 0.277 | 0.190 | 0.061 |
| | K (Critical value) | 3.841 | 3.841 | 3.841 | 3.841 | 3.841 | 3.841 | 3.841 | 3.841 | 3.841 |
| | DF | 1 | 1 | 1 | 1 | 1 | 1 | 1 | 1 | 1 |
| | p-value (one-tailed) | 0.7943 | 0.5820 | 0.4009 | 0.1734 | 0.4512 | 0.3692 | 0.5987 | 0.6632 | 0.8044 |
| 6-17+ | K (Observed value) | 0.722 | 0.722 | 0.029 | 0.260 | 0.029 | 3.494 | 0.097 | 0.080 | 0.787 |
| | K (Critical value) | 3.841 | 3.841 | 3.841 | 3.841 | 3.841 | 3.841 | 3.841 | 3.841 | 3.841 |
| | DF | 1 | 1 | 1 | 1 | 1 | 1 | 1 | 1 | 1 |
| | p-value (one-tailed) | 0.3955 | 0.3955 | 0.8651 | 0.6102 | 0.8651 | 0.0616 | 0.7550 | 0.7768 | 0.3751 |
| 0-6 | K (Observed value) | 0.186 | 0.034 | 0.046 | 0.186 | 0.046 | 0.691 | 0.005 | 0.015 | 0.000 |
| | K (Critical value) | 3.841 | 3.841 | 3.841 | 3.841 | 3.841 | 3.841 | 3.841 | 3.841 | 3.841 |
| | DF | 1 | 1 | 1 | 1 | 1 | 1 | 1 | 1 | 1 |
| | p-value (one-tailed) | 0.6666 | 0.8535 | 0.8294 | 0.6666 | 0.8294 | 0.4059 | 0.9458 | 0.9018 | 1.0000 |
| 0-17+ | K (Observed value) | 0.178 | 0.711 | 0.100 | 0.011 | 0.011 | 0.178 | 0.003 | 0.100 | 0.290 |
| | K (Critical value) | 3.841 | 3.841 | 3.841 | 3.841 | 3.841 | 3.841 | 3.841 | 3.841 | 3.841 |
| | DF | 1 | 1 | 1 | 1 | 1 | 1 | 1 | 1 | 1 |
| | p-value (one-tailed) | 0.6733 | 0.3991 | 0.7518 | 0.9161 | 0.9161 | 0.6733 | 0.9529 | 0.7517 | 0.5901 |

**Table S13**. Kruskal-Wallis tests of fatty acid quantification between ETI treatment time-points and healthy controls.

| | | Fatty acid | | | | | | | | |
|---|---|---|---|---|---|---|---|---|---|---|
| | | Acetic | Propionic | Isobutyric | Butyric | Isovaleric | Valeric | 4-methyl | Hexanoic | Heptanoic |
| 0-HC | K (Observed value) | 0.017 | 0.034 | 0.771 | 0.760 | 0.076 | 14.713 | 1.448 | 5.477 | 12.746 |
| | K (Critical value) | 3.841 | 3.841 | 3.841 | 3.841 | 3.841 | 3.841 | 3.841 | 3.841 | 3.841 |
| | DF | 1 | 1 | 1 | 1 | 1 | 1 | 1 | 1 | 1 |
| | p-value (one-tailed) | 0.8951 | 0.8544 | 0.3798 | 0.3833 | 0.7831 | 0.0001 | 0.2289 | 0.0193 | 0.0004 |
| 3-HC | K (Observed value) | 1.388 | 0.002 | 0.003 | 0.928 | 0.034 | 13.805 | 2.177 | 9.453 | 17.631 |
| | K (Critical value) | 3.841 | 3.841 | 3.841 | 3.841 | 3.841 | 3.841 | 3.841 | 3.841 | 3.841 |
| | DF | 1 | 1 | 1 | 1 | 1 | 1 | 1 | 1 | 1 |
| | p-value (one-tailed) | 0.2387 | 0.9634 | 0.9600 | 0.3353 | 0.8544 | 0.0002 | 0.1400 | 0.0021 | 0.0000 |
| 6-HC | K (Observed value) | 0.852 | 0.021 | 0.754 | 0.517 | 0.589 | 18.621 | 1.019 | 4.056 | 17.806 |
| | K (Critical value) | 3.841 | 3.841 | 3.841 | 3.841 | 3.841 | 3.841 | 3.841 | 3.841 | 3.841 |
| | DF | 1 | 1 | 1 | 1 | 1 | 1 | 1 | 1 | 1 |
| | p-value (one-tailed) | 0.3559 | 0.8856 | 0.3853 | 0.4720 | 0.4430 | 0.0000 | 0.3128 | 0.0440 | 0.0000 |
| 17+-HC | K (Observed value) | 0.375 | 0.034 | 0.576 | 5.952 | 0.010 | 5.952 | 0.146 | 2.143 | 8.109 |
| | K (Critical value) | 3.841 | 3.841 | 3.841 | 3.841 | 3.841 | 3.841 | 3.841 | 3.841 | 3.841 |
| | DF | 1 | 1 | 1 | 1 | 1 | 1 | 1 | 1 | 1 |
| | p-value (one-tailed) | 0.5403 | 0.8544 | 0.4477 | 0.0147 | 0.9223 | 0.0147 | 0.7021 | 0.1432 | 0.0044 |

212

**Table S14**. Kruskal-Wallis tests of fatty acid relative abundance ETI treatment time-points and healthy controls.

| | | Fatty acid | | | | | | | | |
|---|---|---|---|---|---|---|---|---|---|---|
| | | Acetic | Propionic | Isobutyric | Butyric | Isovaleric | Valeric | 4-methyl | Hexanoic | Heptanoic |
| 0-HC | K (Observed value) | 0.157 | 0.004 | 0.352 | 0.526 | 0.000 | 11.309 | 2.968 | 3.915 | 11.373 |
| | K (Critical value) | 3.841 | 3.841 | 3.841 | 3.841 | 3.841 | 3.841 | 3.841 | 3.841 | 3.841 |
| | DF | 1 | 1 | 1 | 1 | 1 | 1 | 1 | 1 | 1 |
| | p-value (one-tailed) | 0.6924 | 0.9474 | 0.5529 | 0.4683 | 1.0000 | 0.0008 | 0.0849 | 0.0478 | 0.0007 |
| 3-HC | K (Observed value) | 0.096 | 0.096 | 0.246 | 0.035 | 1.112 | 8.862 | 1.428 | 5.268 | 11.966 |
| | K (Critical value) | 3.841 | 3.841 | 3.841 | 3.841 | 3.841 | 3.841 | 3.841 | 3.841 | 3.841 |
| | DF | 1 | 1 | 1 | 1 | 1 | 1 | 1 | 1 | 1 |
| | p-value (one-tailed) | 0.7565 | 0.7565 | 0.6198 | 0.8524 | 0.2917 | 0.0029 | 0.2321 | 0.0217 | 0.0005 |
| 6-HC | K (Observed value) | 0.600 | 0.000 | 1.116 | 2.623 | 0.243 | 15.000 | 2.964 | 3.101 | 12.062 |
| | K (Critical value) | 3.841 | 3.841 | 3.841 | 3.841 | 3.841 | 3.841 | 3.841 | 3.841 | 3.841 |
| | DF | 1 | 1 | 1 | 1 | 1 | 1 | 1 | 1 | 1 |
| | p-value (one-tailed) | 0.4386 | 1.0000 | 0.2908 | 0.1053 | 0.6221 | 0.0001 | 0.0851 | 0.0782 | 0.0005 |
| 17+-HC | K (Observed value) | 0.735 | 0.735 | 2.535 | 1.815 | 0.015 | 9.375 | 0.462 | 1.815 | 4.343 |
| | K (Critical value) | 3.841 | 3.841 | 3.841 | 3.841 | 3.841 | 3.841 | 3.841 | 3.841 | 3.841 |
| | DF | 1 | 1 | 1 | 1 | 1 | 1 | 1 | 1 | 1 |
| | p-value (one-tailed) | 0.3913 | 0.3913 | 0.1113 | 0.1779 | 0.9025 | 0.0022 | 0.4967 | 0.1779 | 0.0372 |

213

**Table S15**. ANOSIM summary statistics of fatty acid relative abundances across ETI treatment time-points and compared with healthy control participants.

| | Across ETI Therapy | | | Between ETI & Healthy Controls | |
|---|---|---|---|---|---|
| 0-3 | R | -0.0061 | 0-HC | R | 0.0794 |
| | p (same) | 0.4428 | | p (same) | 0.105 |
| | Bonferroni-corrected p value | 0.4516 | | Bonferroni-corrected p value | 0.1067 |
| | Permutations | 9999 | | Permutations | 9999 |
| 3-6 | R | -0.0188 | 3-HC | R | -0.0013 |
| | p (same) | 0.5393 | | p (same) | 0.4222 |
| | Bonferroni-corrected p value | 0.5417 | | Bonferroni-corrected p value | 0.4367 |
| | Permutations | 9999 | | Permutations | 9999 |
| 6-17+ | R | -0.0624 | 6-HC | R | 0.1502 |
| | p (same) | 0.6692 | | p (same) | 0.0393 |
| | Bonferroni-corrected p value | 0.6744 | | Bonferroni-corrected p value | 0.0407 |
| | Permutations | 9999 | | Permutations | 9999 |
| 0-6 | R | -0.0475 | 17+-HC | R | 0.2480 |
| | p (same) | 0.8064 | | p (same) | 0.0467 |
| | Bonferroni-corrected p value | 0.8122 | | Bonferroni-corrected p value | 0.047 |
| | Permutations | 9999 | | Permutations | 9999 |
| 0-17+ | R | -0.1939 | | | |
| | p (same) | 0.9724 | | | |
| | Bonferroni-corrected p value | 0.9757 | | | |
| | Permutations | 9999 | | | |

**Table S16.** Similarity of percentage (SIMPER) analysis of fatty acid compositional dissimilarity between healthy controls and pwCF following 6 months and 17+ months (extended) ETI therapy.

| | % Relative abundance | | | | |
|---|---|---|---|---|---|
| Fatty acid | ETI 6 Months | Healthy Controls | Av. Dissimilarity | % Contribution | Cumulative (%) |
| Butyric | 27.3 | 21.4 | 4.767 | 28.24 | 28.24 |
| Acetic | 49.7 | 50.6 | 3.931 | 23.28 | 51.52 |
| Propionic | 17.5 | 17.5 | 3.236 | 19.17 | 70.69 |
| Valeric | 0.276 | 3.15 | 1.438 | 8.517 | 79.2 |
| Hexanoic | 0.678 | 2.19 | 1.177 | 6.974 | 86.18 |
| Isobutyric | 2.2 | 2.54 | 1.112 | 6.586 | 92.76 |
| Isovaleric | 2.4 | 2.21 | 1.048 | 6.209 | 98.97 |
| Heptanoic | 0.0045 | 0.305 | 0.1502 | 0.8899 | 99.86 |
| 4-methyl | 0.0209 | 0.049 | 0.02314 | 0.137 | 100 |
| **Total dissimilarity** | | | **16.88** | | |

| | % Relative abundance | | | | |
|---|---|---|---|---|---|
| Fatty acid | ETI 17+ | Healthy Controls | Av. Dissimilarity | % Contribution | Cumulative (%) |
| Acetic | 46.9 | 50.6 | 4.823 | 28.73 | 28.73 |
| Butyric | 27.5 | 21.4 | 4.329 | 25.79 | 54.52 |
| Propionic | 21 | 17.5 | 3.427 | 20.41 | 74.93 |
| Valeric | 0.458 | 3.15 | 1.347 | 8.023 | 82.95 |
| Hexanoic | 0.416 | 2.19 | 1.065 | 6.341 | 89.29 |
| Isobutyric | 1.45 | 2.54 | 0.8098 | 4.824 | 94.12 |
| Isovaleric | 2.17 | 2.21 | 0.8068 | 4.806 | 98.92 |
| Heptanoic | 0.01 | 0.305 | 0.1491 | 0.8881 | 99.81 |
| 4-methyl | 0.05 | 0.049 | 0.0321 | 0.1912 | 100 |
| **Total dissimilarity** | | | **16.79** | | |

**Table S17**. RDA to explain percent variation in microbiota from faecal relative SCFA abundance.

| SCFA | Var. Exp (%) | pseudo-*F* | *P* (adj) |
|---|---|---|---|
| Valeric | 4 | 2 | 0.002 |
| Propionic | 4 | 2 | 0.002 |
| Butyric | 2.7 | 1.4 | 0.018 |
| Total | **10.7** | | |

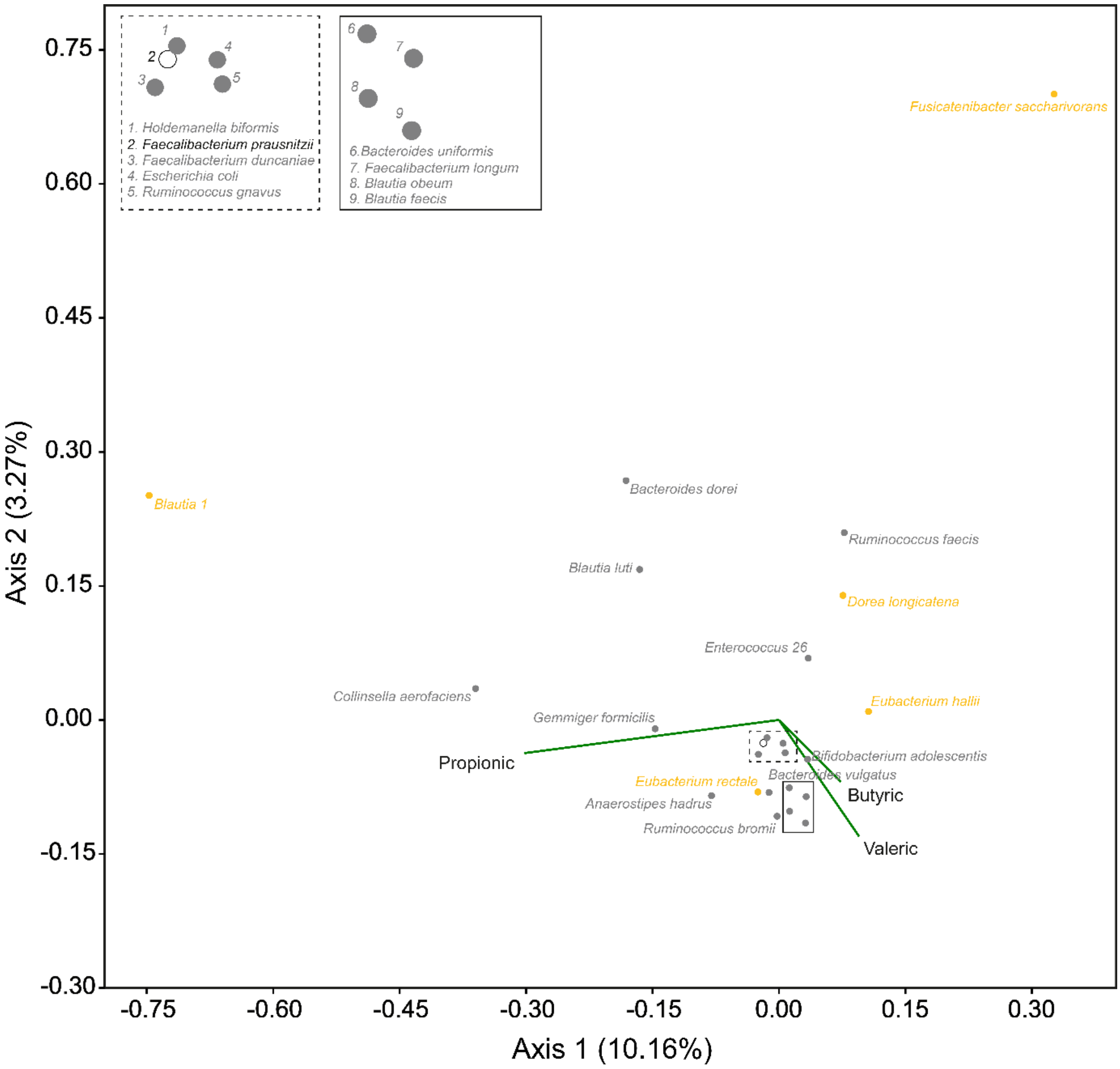

**Figure S4.** Faecal SCFA redundancy analysis species biplots for the whole microbiota. The 24 taxa contributing most to the dissimilarity (cumulatively > 50%) between healthy control and pwCF samples following 17+ months (extended) ETI therapy from the SIMPER analysis (Table 2) are shown independently of the total number of ASVs identified (531). Orange points represent taxa that were identified as core for the pwCF group following 17+ (extended) ETI therapy, grey points are satellite, and the white (black stroke) points represent taxa that were absent. Biplot lines depict SCFA that significantly explained total variation in taxa relative abundance within whole microbiota analysis at the $p \leq 0.05$ level (Table S17). Species plots depict the strength of explanation provided by the given clinical variables, with taxa shown in the same direction of a SCFA considered to have a higher value than those that are not. The percentage of microbiota variation explained by each axis is given in parentheses.

**Table S18**. Summary statistics for paired t-test PAC-SYM results between baseline and ETI treatment

| | | |
|---|---|---|
| 3 | Difference | -0.339 |
| | t (Observed value) | -0.398 |
| | \|t\| (Critical value) | 2.160 |
| | DF | 13 |
| | p-value (Two-tailed) | 0.697 |
| 6 | Difference | -1.750 |
| | t (Observed value) | -1.923 |
| | \|t\| (Critical value) | 2.179 |
| | DF | 12 |
| | p-value (Two-tailed) | 0.079 |
| 17+ | Difference | -1.429 |
| | t (Observed value) | -1.000 |
| | \|t\| (Critical value) | 2.447 |
| | DF | 6 |
| | p-value (Two-tailed) | 0.356 |

228

**Table S19**. Kruskal-Wallis summary statistics for gut function MRI metrics between baseline and 17+ (extended) ETI samples.

| | | |
|---|---|---|
| OCTT | K (Observed value) | 0.040 |
| | K (Critical value) | 3.841 |
| | DF | 1 |
| | p-value (one-tailed) | 0.842 |

| | | |
|---|---|---|
| SBWC | K (Observed value) | 3.438 |
| | K (Critical value) | 3.841 |
| | DF | 1 |
| | p-value (one-tailed) | 0.064 |
